# The effect of herpes zoster vaccination on the occurrence of deaths due to dementia in England and Wales

**DOI:** 10.1101/2023.09.08.23295225

**Authors:** Felix Michalik, Min Xie, Markus Eyting, Simon Heß, Seunghun Chung, Pascal Geldsetzer

## Abstract

**Background:** The United Kingdom (UK) has used date of birth-based eligibility rules for live-attenuated herpes zoster (HZ) vaccination that have led to large differences in HZ vaccination coverage between individuals who differed in their age by merely a few days. Using this unique natural randomization, we have recently provided evidence from Welsh electronic health record data that HZ vaccination caused a reduction in new dementia diagnoses over a seven-year period. Based on this, we hypothesized that HZ vaccination may have slowed the dementia disease process more generally and, thus, already reduced deaths with dementia as their underlying cause even though the UK’s HZ vaccination program commenced as recently as September 2013. Using country-wide death certificate data for England and Wales, this study, therefore, aimed to determine whether eligibility for HZ vaccination caused a reduction in deaths due to dementia over a nine-year follow-up period.

**Methods:** Adults who had their 80^th^ birthday shortly before September 1 2013 were ineligible for HZ vaccination in the UK’s National Health Service and remained ineligible for life, whereas those who had their 80^th^ birthday shortly after September 1 2013 (i.e., born on or after September 2 1933) were eligible for one year. Akin to a randomized trial, this date-of-birth threshold generated birth cohorts who are likely exchangeable in observed and unobserved characteristics except for a small difference in age and a large difference in HZ vaccination uptake. We used country-wide data from death certificates in England and Wales on underlying causes of death from September 1 2004 to August 31 2022 by ICD-10 code and month of birth. Our analysis compared the percentage of the population with a death due to dementia among the month-of-birth cohorts around the September 2 1933 eligibility threshold using a regression discontinuity design. The primary analyses used the maximal available follow-up period of nine years.

**Results:** The study population included 5,077,426 adults born between September 1 1925 and August 31 1941 who were alive at the start of the HZ vaccination program. The month-of-birth cohorts around the September 2 1933 eligibility cutoff were well balanced in their occurrence of all-cause and cause-specific deaths (including deaths due to dementia) prior to the start of the vaccination program. We estimated that over a nine-year follow-up period, eligibility for HZ vaccination reduced the percentage of the population with a death due to dementia by 0.38 (95% CI: 0.08 to 0.68, p=0.012) percentage points, corresponding to a relative reduction of 4.8%. As in our prior analysis, this effect was stronger among women (−0.62 [95% CI: −1.06 to −0.19] percentage points, p=0.004) than among men (−0.11 [95% CI: −0.51 to 0.28] percentage points, p=0.574). The reduction in deaths due to dementia likely resulted in an increase in remaining life expectancy because we found that HZ vaccination eligibility reduced all-cause mortality but had no effect on deaths not due to dementia. An effect on deaths due to dementia at the September 2 date-of-birth eligibility threshold existed only since the year in which the HZ vaccination program was implemented.

**Conclusions:** Our findings indicate that HZ vaccination improved cognitive function at a fairly advanced stage of the dementia disease process because most individuals whose underlying cause of death was dementia during our nine-year follow-up period were likely already living with dementia at the start of the HZ vaccination program. By using a different population, type of data, and outcome than our prior study in Welsh electronic health record data, this analysis adds to the evidence base that HZ vaccination slows, or potentially even prevents, the natural history of dementia.

## Background

Partly owing to the increasing recognition of long-term neurological sequelae from SARS-CoV-2 infections(1), as well as the accumulating evidence on the Epstein-Barr-Virus as a causative agent in the development of Multiple Sclerosis(2), the possibility that neurotropic viruses may play a role in the pathogenesis of a subset of dementia cases has recently garnered increasing attention(3–5). The largest body of evidence for a viral etiology of dementia exists for herpesviruses(3–7). At present, a phase 2 proof-of-concept trial is being funded by the US National Institute on Aging, which studies the impact of an antiviral drug on the cognitive and functional ability of individuals with mild Alzheimer’s dementia(8). In addition to the possibility of a viral etiology of dementia, there is increasing consensus that the immune system and neuroinflammation play a critical role in the development of dementia(9), and that live-attenuated vaccines appear to induce broader immune mechanisms that have important off-target health effects(10–12). These off-target effects of vaccines are often far more pronounced among female than male individuals(11).

Our group has recently exploited a natural randomization in Wales, along with country-wide electronic health record data, to provide evidence that live-attenuated herpes zoster (HZ) vaccination caused a reduction in new dementia diagnoses over a five- to eight-year follow-up period(13). Specifically, we took advantage of the fact that, starting on September 1 2013, adults born on or after September 2 1933 were eligible for one year for live-attenuated HZ vaccination (ZOSTAVAX [Merck]) in the United Kingdom’s National Health Service (NHS) whereas those born earlier never became eligible. Because there is no plausible reason why those born just before versus just after September 2 1933 would systematically differ from each other, this approach compared – just like in a randomized trial – two groups that are in expectation identical to each other on observed and unobserved characteristics except for a large difference in their probability of receiving the intervention of interest. Our unique approach is, thus, not subject to the fundamental concern in the existing body of epidemiological evidence(14) that those who opt to be vaccinated differ from those who do not (i.e., selection bias) in a myriad of characteristics that are difficult or impossible to measure(15).

We hypothesized that if HZ vaccination reduces new dementia diagnoses, then it may slow the natural history of dementia more generally and, thus, also reduce the occurrence of deaths due to dementia among individuals who already have a degree of dementia. Employing the identical natural randomization approach as our study in Welsh electronic health record data(13), the primary aim of this study was, therefore, to determine the effect of being eligible for HZ vaccination on the occurrence of deaths due to dementia during the nine years following the start of the HZ vaccination program. Conditional on the primary aim of the study identifying a significant reduction in deaths due to dementia from HZ vaccination eligibility, our secondary aim was to determine if being eligible for HZ vaccination also reduced all-cause mortality and, thus, increased remaining life expectancy.

Individuals whose death during our nine-year follow-up period was recorded as having dementia as its underlying cause likely already had a degree of dementia at the beginning of the follow-up period(16). This study, thus, provides new insights beyond those gained from our previous analysis in Welsh electronic health record data because it determines the effect of HZ vaccination on dementia progression at a different, later stage of the dementia disease process. An additional contribution of this study is that it examines if the findings of our prior analysis hold true when studying a different population (England’s population accounts for approximately 95% of England’s and Wales’s combined population(17)), type of data (death certificates as opposed to electronic health records), and outcome.

## Methods

### Data sources

For the period of September 1 2004 to August 31 2022, the United Kingdom’s (UK’s) Office of National Statistics (ONS) provided us with six-monthly numbers of all deaths that took place in England and Wales by month of birth and sex for adults born between September 1 1925 and August 31 1941. In addition to all-cause mortality, death numbers (as sourced from death certificates(18)) were provided separately for each of the 68 leading underlying causes of death in England and Wales (using ONS’s 2016 revised version(19)). More detailed methodological information on mortality statistics in the UK is available elsewhere(18). We obtained data on the size of the population in England and Wales as of March 27 2011 by month of birth from the 2011 UK Census(20). We then calculated population numbers (by month of birth) for each six-monthly period from September 1 2004 to August 31 2022 by subtracting or adding the total number of deaths in each month-of-birth cohort from the 2011 month-of-birth population numbers in the census data.

### Outcome definition

Our primary outcome of interest was the probability of dying from dementia after the start of the HZ vaccination program. Because our data was aggregated at the level of a birth-month cohort, we defined our primary outcome as the percentage of the population alive on September 1 2013 (the start date of the HZ vaccination program) whose underlying cause of death was dementia during a nine-year follow-up period. Given the neuropathological overlap between dementia types(21–23), the difficulty in distinguishing dementia types clinically(24), and the reduction in statistical power when examining a rarer outcome, dementia was defined as dementia of any type in our primary analyses. In exploratory analyses, we show results separately for each of the three types of dementia in our data (Alzheimer’s disease, vascular dementia, and dementia of unspecified type). Our secondary outcome was the percent of the population alive on September 1 2013 who died from any cause during the nine-year follow-up period.

### Exposure definition

The exposure was eligibility for HZ vaccination in the NHS as determined by an individual’s date of birth(25). Adults born between September 2 1933 and September 1 1934 became eligible for HZ vaccination on September 1 2013. Eligibility was then gradually extended to younger date-of-birth cohorts on an annual basis (details are provided in **Text S1**); older cohorts never became eligible. Those who became eligible for HZ vaccination on September 1 2013, thus, remained eligible for one year. We have previously shown in country-wide electronic health record data from Wales that there was an abrupt increase at the September 2 1933 date-of-birth eligibility threshold in the probability of ever having received the HZ vaccine from 0.01% to 47.2%(13).

### Statistical analysis

#### General analytical approach

We tested for differences in our outcomes across the September 2 1933 date-of-birth eligibility threshold for HZ vaccination using regression discontinuity analysis, which is a well-established approach for causal effect estimation(26). This analysis assumed that individuals born just around the September 2 1933 threshold are exchangeable with each other in both observed and unobserved characteristics that could influence deaths due to dementia. In our view, this assumption is justified because i) it appears unlikely that the exact date of birth of September 2 1933 was used by a different relevant intervention as its eligibility criterion, and we additionally provide evidence against this possibility (see the “Testing for confounding” section below); ii) it is impossible that parents in the 1930’s could have planned the timing of their births according to an HZ vaccination program that was implemented in 2013; iii) we have demonstrated in country-wide electronic health record data from Wales that individuals are well balanced in their observable characteristics across the September 2 1933 threshold(13); and iv) we show that the month-of-birth cohorts just around the September 2 1933 threshold are well balanced in their probability of deaths from any cause and from each of the ten most common causes of death in the nine years preceding the HZ vaccination program.

All regression equations used in our analyses are shown in **Supplement Text S2**. Our regression discontinuity analysis estimated the difference in the probability of dying due to dementia between individuals who were born just on either side of the date-of-birth eligibility threshold. Thus, our parameter of interest was the discontinuity in the outcome, conditional on month of birth, at the September 2 1933 threshold. As per recommended practice for regression discontinuity analyses(27–29), we estimated this parameter using local linear regression with triangular kernel weights (giving, linearly, more weight to those observations at the birth-month level that were closer to the threshold) within a mean squared-error (MSE) optimal bandwidth. The bandwidth restricts the effective sample to observations that are sufficiently close to the threshold. The MSE criterion is used as an objective criterion to trade off between precision and possible misspecification. The effective sample size for each regression are stated underneath the corresponding figures in the main manuscript and Supplement. Because our assignment variable (month of birth) was discrete rather than continuous, we computed so-called “honest” confidence intervals based on the approach by Armstrong and Kolesar(30,31). This approach guards against potential vulnerability to model misspecification and resulting under-coverage of confidence intervals computed with more standard methods(30). These honest confidence intervals are conservative in the sense that they have good coverage properties irrespective of whether the functional form in the regression discontinuity analysis is misspecified, provided that the true functional form falls within a certain class of functions. For this class, we considered a function class defined by bounds on the second derivative of the conditional expectation function mapping date of birth to the probability of dying from dementia. The bounds that we use vary across regressions, are selected following Armstrong and Kolesar(31), and are provided in **Tables S2** and **S3** for each of our analyses. All confidence intervals and p-values in the results section were estimated using the approach described in this section. Given their derivation, our honest confidence intervals and p-values should be interpreted as tending to over-rather than underestimate the statistical uncertainty of our estimates.

Regression discontinuity analyses generally yield absolute effect estimates as they are based on estimation via local linear regression. We additionally calculated relative effect estimates by dividing the regression discontinuity estimate of the absolute effect by the predicted value of the outcome just left (i.e., among those who were ineligible for HZ vaccination) of the threshold.

#### Exchangeability across the September 2 1933 threshold

We conducted balance tests to provide empirical evidence that individuals just around the September 2 1933 threshold were indeed exchangeable with each other. Specifically, we implemented the identical regression discontinuity analysis as we used for our outcome analyses except that we used the percentage of the population that died from any cause during the nine years *preceding* the start of the HZ vaccination program (i.e., the period of September 1 2004 to August 31 2013) as a summary measure of mortality risk. In addition, we conducted these tests for each of the ten most common causes of death in our data. We show these balance tests for the whole population and separately for women and men, for a total of 33 tests.

#### Effect on deaths due to dementia and all-cause mortality

To test for the effect of eligibility for HZ vaccination on deaths due to dementia and all-cause mortality, we used a follow-up period of nine years (the longest follow-up period available in our data) in our primary analyses. We, however, also show all results when using instead follow-up periods of 6.0, 6.5, 7.0, 7.5, 8.0, 8.5, and 9.0 years. Similarly, while we used a grace period (the time period since September 1 2013 after which follow-up time is considered to start to allow for the time needed for HZ vaccination eligibility to begin affecting our outcomes) of zero months in our primary analyses, we also show all results when using grace periods of six, 12, 18, 24, 30, and 36 months.

#### Attributing effects on all-cause mortality to effects on deaths due to dementia

Our hypothesis was that eligibility for HZ vaccination reduced deaths due to dementia and, in doing so, may also have decreased all-cause mortality. We, thus, investigated whether any observed effects on all-cause mortality could be attributed to effects on deaths due to dementia. For this analysis, we opted against simply comparing the range of estimates covered by our 95% confidence intervals for the effect of HZ vaccination eligibility on deaths due to dementia versus all-cause mortality, because deaths due to dementia as recorded on death certificates are known to underestimate the number of deaths attributable to dementia(32,33). Instead, we used two strategies that we anticipated would be less vulnerable to this underestimation of deaths attributable to dementia. First, using the same regression discontinuity models as for our primary analyses, we estimated the effect of HZ vaccination eligibility on deaths that did not have dementia as their underlying cause (“non-dementia deaths”). If we were to find significant effects (in the same direction) for deaths due to dementia and all-cause mortality but not for non-dementia deaths, this analysis would support the assumption that all-cause mortality effects could be attributed to effects on deaths due to dementia. Second, we used a competing risk survival model to more formally consider the competing risk between deaths due to dementia and non-dementia deaths. To do so, we used our population number estimates for September 1 2013 (for individuals born between September 1 1925 and August 31 1941) by month of birth and gender as a synthetic cohort by constructing an individual-level dataset from our month of birth-level aggregated data. Our mortality data by month of birth and gender was provided for each six-monthly period between September 1 2013 and the end of our nine-year follow-up period. We censored individuals’ follow-up time in each month-of-birth cohort at the end of the six-month interval in which a person died or (for those who did not die during the follow-up period) at the end of the follow-up period. We then used a Cox regression model to estimate the hazard rate separately for each of two outcomes: deaths due to dementia and non-dementia deaths. The regression was implemented with observations from the same bandwidth around the date-of-birth eligibility threshold that was considered MSE-optimal in our primary regression discontinuity analysis with deaths due to dementia as the outcome. Finally, we obtained the cumulative incidence functions for deaths due to dementia by HZ vaccination eligibility status, and estimated the standardized absolute risk difference and risk ratio (RR) for the effect of being eligible for HZ vaccination on deaths due to dementia(34,35). Our Cox regression approach assumed proportional hazards and, to be able to use the G-formula, that our outcome model was correctly specified. The equations for these regressions are detailed in **Text S2**.

#### Effect heterogeneity by gender

We additionally implemented our primary analysis separately by gender for the following reasons: i) the observation in our previous analysis in electronic health record data from Wales that the effect of HZ vaccination on dementia incidence was stronger among women than men(13); ii) the evidence that shingles occurs more commonly among women(36,37); iii) the growing body of evidence suggesting that there may be differences in the pathogenesis of dementia between women and men(38–40); iv) the observation that off-target effects of vaccines are generally far more pronounced among female than male individuals(11); and v) sex differences in the immunological response to vaccines more broadly(41). We formally tested for the statistical significance of any effect heterogeneity by gender by estimating a regression discontinuity design regression equation that was fully interacted with a binary gender variable. Specifically, we used the MSE-optimal bandwidth by gender from our primary regression model and then estimated the effect difference between women and men via triangularly weighted least-squared regressions. The regression equations for this test are provided in **Text S2**. We also implemented these tests separately by type of dementia (Alzheimer’s disease, vascular dementia, and dementia of unspecified type).

#### Testing for confounding

The key strength of our regression discontinuity approach is that confounding is only possible if there is a confounding variable that changes abruptly at the September 2 1933 date-of-birth threshold. Such a confounder would be an intervention or policy (e.g., another vaccination program) that used the exact same date-of-birth eligibility threshold of September 2 1933 as its eligibility criterion. We tested for the existence of such an intervention in three ways. First, because such an intervention is unlikely to solely affect deaths due to dementia, we implemented the same analysis as for deaths due to dementia and all-cause mortality as an outcome for each of the ten most common causes of death in our data during our nine-year follow-up period. We henceforth refer to these analyses as negative control outcome analyses(42). Second, we implemented the identical analysis as for September 1 2013 (the actual date on which the HZ vaccination program started) for September 1 of each of the six years prior to 2013. For example, when shifting the start date of the vaccination program to 2012, we compared individuals around the September 2 1932 eligibility threshold with the follow-up period starting on September 1 2012. If there was an intervention or policy that used a September 2 date-of-birth eligibility rule, then we would expect to observe an effect of the September 2 threshold on deaths due to dementia in years other than only the year in which the HZ vaccination program started. Third, we implemented the identical month of birth-cohort comparison (i.e., comparing cohorts just around the September 2 1933 date-of-birth threshold) as in our primary analyses, but used deaths due to dementia and all-cause mortality in the nine years *preceding* September 1 2013 as the outcome. If an intervention that had a sizeable effect on deaths due to dementia used September 2 1933 as its date-of-birth eligibility criterion and the intervention was implemented before 2013, then it would likely affect deaths due to dementia already prior to the start of the HZ vaccination program. As with our balance checks, we conducted all tests for confounding described in this section for the whole population and separately for women and men. In total, we therefore performed 69 tests for confounding.

#### Robustness checks

In addition to using different follow-up and grace periods (as detailed above), we used local squared instead of local linear regression to verify the robustness of our results to a more flexible functional form. Following current best practice for regression discontinuity analyses(27), we did not use global polynomial regressions of higher order. We further verified whether our point estimates remained similar when using different bandwidths (12, 18, and 24 months) than the MSE-optimal bandwidth.

#### Additional analyses

Our negative control outcome analyses identified a significant effect of being eligible for HZ vaccination on deaths due to cerebrovascular disease. Transient ischemic attacks and strokes as a result of varicella zoster virus-induced vasculopathy that affects the intracerebral arteries is a well-recognized complication of shingles(43–45). Because this complication of shingles most often occurs within a few months of a shingles episode and because the protection for shingles from live-attenuated HZ vaccination wanes over time(46,47), we hypothesized that the relative effect of HZ vaccination eligibility on deaths due to cerebrovascular disease would decline over the duration of our follow-up period. To investigate this hypothesis, we plotted both the absolute and relative effect of HZ vaccination eligibility on deaths due to cerebrovascular disease across follow-up periods ranging from one to nine years in six-month increments. We conducted the same robustness checks (described above) as for deaths due to dementia and all-cause mortality also for deaths due to cerebrovascular disease.

## Results

### Study population

Our study population consisted of a total of 5,077,426 individuals (2,824,529 women and 2,252,897 men) born between September 1 1925 and August 31 1941 who were estimated to be alive on September 1 2013 (**Table 1**). **Figure S1** disaggregates this study population by month of birth. In the nine years preceding September 1 2013 (the start of the HZ vaccination program), 1,721,374 (792,224 women and 929,150 men) individuals born between September 1 1925 and August 31 1941 had died, with the most common underlying causes of death being cancers other than lung cancer, ischemic heart disease, lung cancer, cerebrovascular disease, chronic lower respiratory diseases, other circulatory diseases, dementia, acute respiratory infections, heart failure and hypertensive diseases, and diseases of the urinary system. Table 1 displays the number of deaths in the nine years preceding September 1 2013 for each of the ten most common underlying causes of death during the follow-up period and **Table S1** further disaggregates these causes by each cause of death in our dataset. Table S1 also lists the ICD-10 codes that correspond to each cause of death in our data.

**Table 1.** Characteristics of the study population^1^.

| Variable | Total |  | Women |  | Men |  |
| --- | --- | --- | --- | --- | --- | --- |
|  | N | % | N | % | N | % |
| Study population in Sept 2013 | 5,077,426 | 100 | 2,824,529 | 55.6 | 2,252,897 | 44.4 |
| Study population in Sept 2004 | 6,798,800 | 100 | 3,616,753 | 53.2 | 3,182,047 | 46.8 |
| *Deaths from Sept 1 2004 to Aug 31 2013<sup>2</sup>* |  |  |  |  |  |  |
| All-cause mortality | 1,721,374 | 25.5 | 792,224 | 22.1 | 929,150 | 29.5 |
| Other cancers<sup>3</sup> | 369,757 | 5.5 | 166,337 | 4.6 | 203,420 | 6.5 |
| Ischemic heart disease | 276,903 | 4.1 | 102,096 | 2.8 | 174,807 | 5.5 |
| Lung cancer<sup>4</sup> | 140,304 | 2.1 | 58,791 | 1.6 | 81,513 | 2.6 |
| Cerebrovascular diseases | 133,599 | 2.0 | 69,659 | 1.9 | 63,940 | 2.0 |
| Chronic lower respiratory diseases | 120,233 | 1.8 | 57,567 | 1.6 | 62,666 | 2.0 |
| Other circulatory diseases<sup>5</sup> | 78,750 | 1.2 | 36,515 | 1.0 | 42,235 | 1.3 |
| Dementia | 75,770 | 1.1 | 44,893 | 1.3 | 30,877 | 1.0 |
| Acute respiratory infections<sup>6</sup> | 71,240 | 1.1 | 35,267 | 1.0 | 35,973 | 1.1 |
| Heart failure and hypertensive diseases | 38,314 | 0.6 | 19,153 | 0.5 | 19,161 | 0.6 |
| Diseases of the urinary system | 30,154 | 0.5 | 15,952 | 0.4 | 14,202 | 0.5 |

<sup>1</sup> The ICD-10 codes for all causes of death in our data are shown in Table S1.
<sup>2</sup> The percentages for the deaths from September 1 2004 to August 31 2013 were obtained by dividing N by the population in September 2004.
<sup>3</sup> Defined as cancers other than cancers of the trachea, bronchus, and lung.
<sup>4</sup> Defined as cancers of the trachea, bronchus, and lung.
<sup>5</sup> Other circulatory diseases were defined as ICD-10 codes I05–I09, I26–I28, I34–I38, I42, I46, I47–I49, I70, and I71.
<sup>6</sup> Acute respiratory infections were defined as ICD-10 codes J00–J06, J09–J18, and J20–J22.
Abbreviations: Sept=September; Aug=August; HF=heart failure

### Exchangeability across the September 2 1933 threshold

There were no significant differences across the September 2 1933 date-of-birth eligibility threshold in the percentage of the population alive on September 1 2004 that died from any cause, nor from each of the top ten causes of death, in the nine years prior to the start of the HZ vaccination program (**Figure 1**). This also held true when analyzing men only (**Figure S2**). Among women, one cause of death (cancer other than lung cancer) showed a significant difference (−0.17 [95% CI: −0.34 to −0.01] percentage points, p=0.040) (**Figure S3**).

**Figure 1.**
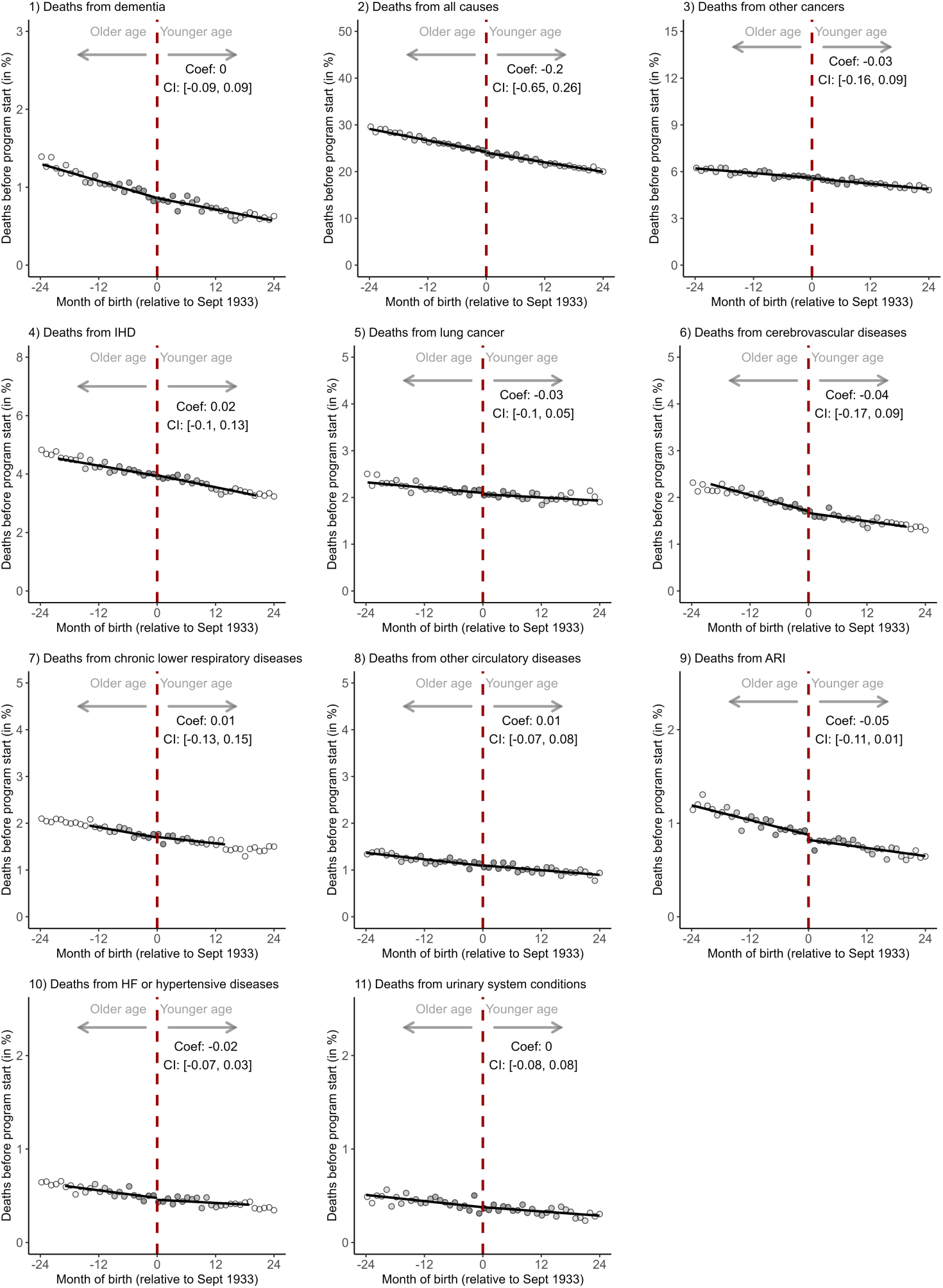
Balance tests across the date-of-birth eligibility threshold for HZ vaccination.^1,2,3,4,5,6,7,8^. ^1^ The y-axis shows the percent of the population in each month-of-birth cohort alive on September 1 2004 that died from the given cause in the nine years preceding the start of the HZ vaccination program. ^2^ The grey shading of the dots is in proportion to the weight that observations from this birth-month received in the analysis. ^3^ The linear regression lines are drawn only in the mean-squared-error optimal bandwidth. ^4^ Other cancers were defined as cancers other than cancers of the trachea, bronchus, and lung. ^5^ Lung cancer encompassed cancers of the trachea, bronchus, and lung. ^6^ Other circulatory diseases were defined as ICD-10 codes I05–I09, I26–I28, I34–I38, I42, I46, I47–I49, I70, and I71. ^7^ Acute respiratory infections were defined as ICD-10 codes J00–J06, J09–J18, and J20–J22. ^8^ The ICD-10 codes to define each cause of death are shown in Table S1. Abbreviations: Coef=coefficient; CI=95% confidence interval; IHD=ischemic heart diseases; ARI=acute respiratory infections; HF=heart failure

### Effect of being eligible for HZ vaccination on deaths due to dementia

A total of 373,548 individuals (234,502 women and 139,046 men) born between September 1 1925 and August 31 1941 and who were alive on the start date of the HZ vaccination program died due to dementia during the nine-year follow-up period. Being eligible for HZ vaccination reduced the percent of this population that died due to dementia by an estimated 0.38 (95% CI: 0.08 to 0.68, p=0.012) percentage points (**Figure 2**), corresponding to a relative reduction of 4.8%. The absolute effect of being eligible for HZ vaccination on the proportion of the population that died due to dementia increased with an increasing follow-up period and reached statistical significance at 8.5 and 9.0 years of follow-up. The relative effect also increased with an increasing follow-up period, from 2.7% at six years to 4.8% at nine years. The effect was consistent across grace periods ranging from zero to 36 months. None of the effect estimates reached statistical significance when examining deaths due to Alzheimer’s disease, vascular dementia, and dementia of unspecified type separately (**Table S2**).

**Figure 2.**
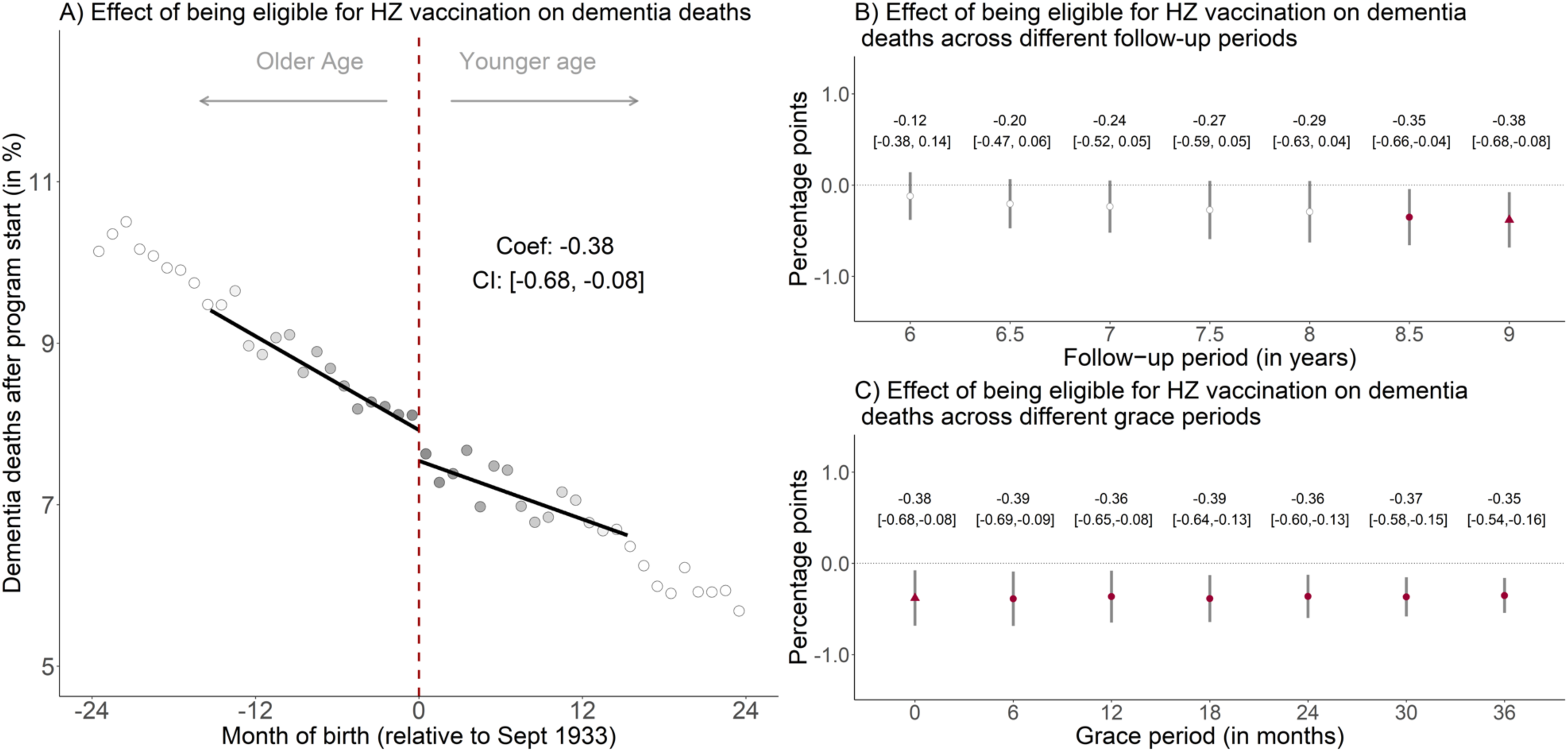
The effect of being eligible for HZ vaccination on the percent of the population alive on September 1 2013 that died due to dementia.^1,2,3,4,5,6,7^. ^1^ Triangles (rather than points) depict our primary specification. ^2^ Red (as opposed to white) fillings denote statistical significance (p<0.05). ^3^ The grey shading of the dots is in proportion to the weight that observations from this birth-month received in the analysis. ^4^ With “grace periods” we refer to time periods since September 1 2013 after which follow-up time is considered to begin to allow for the time needed for HZ vaccination eligibility to begin affecting the outcome. ^5^ Grey vertical bars depict 95% confidence intervals. ^6^ In Panel A, the sample size in the mean squared error-optimal bandwidth is 823,548 adults. ^7^ The linear regression lines are drawn only in the mean squared error-optimal bandwidth. Abbreviations: Coef=coefficient; CI=95% confidence interval; HZ=herpes zoster; Sept=September

### Effect heterogeneity between women and men

Among women, being eligible for HZ vaccination reduced the percentage alive on September 1 2013 that died due to dementia by 0.62 (95% CI: 0.19 to 1.06, p=0.004) percentage points over our nine-year follow-up period (**Figure 3**), corresponding to a relative reduction of 7.0%. The absolute effect size increased with an increasing follow-up period and became statistically significant after 6.5 years of follow-up. The effect was consistent across grace periods from zero to 36 months. Among men, there was no significant effect (−0.11 [95% CI: −0.51 to 0.28] percentage points, p=0.574). The effect heterogeneity by gender was statistically significant in our interaction tests (**Table S3**). None of the interaction tests reached significance when examining these effects separately for each type of dementia (Table S3).

**Figure 3.**
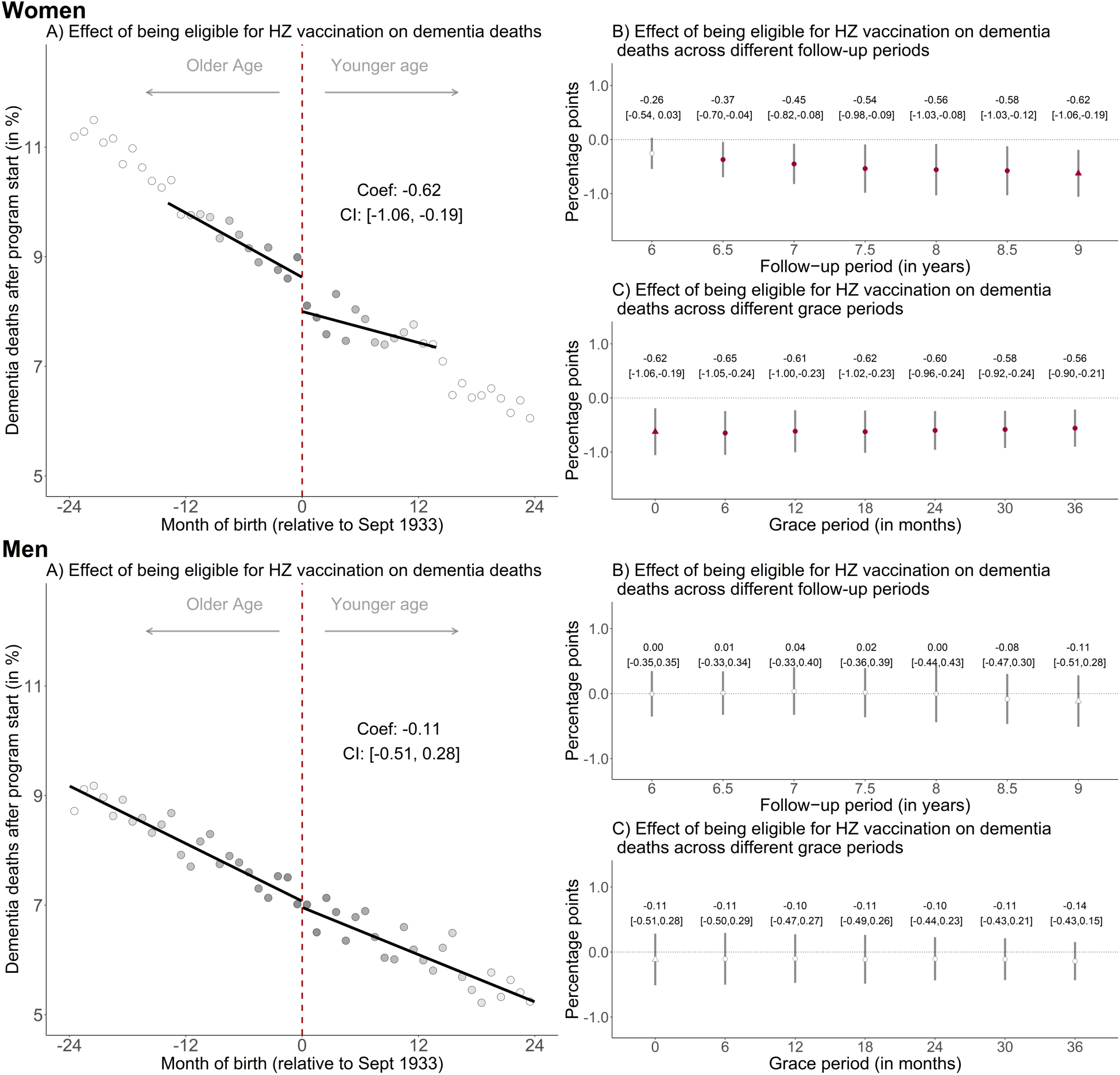
The effect of being eligible for HZ vaccination on the percent of the population alive on September 1 2013 that died due to dementia, by sex.^1,2,3,4,5,6,7^. ^1^ Triangles (rather than points) depict our primary specification. ^2^ Red (as opposed to white) fillings denote statistical significance (p<0.05). ^3^ The grey shading of the dots is in proportion to the weight that observations from this birth-month received in the analysis. ^4^ With “grace periods” we refer to time periods since September 1 2013 after which follow-up time is considered to begin to allow for the time needed for HZ vaccination eligibility to begin affecting the outcome. ^5^ Grey vertical bars depict 95% confidence intervals. ^6^ In Panel A, the sample size in the mean squared error-optimal bandwidth is 398,376 and 585,157 for women and men, respectively. ^7^ The linear regression lines are drawn only in the mean squared error-optimal bandwidth. Abbreviations: Coef=coefficient; CI=95% confidence interval; HZ=herpes zoster; Sept=September

### Effect of being eligible for HZ vaccination on all-cause mortality

Of the 5,077,426 individuals in our study population who were alive on September 1 2013, 2,436,067 individuals (1,262,588 women and 1,173,479 men) died during the nine-year follow-up period. Eligibility for HZ vaccination decreased the percent of this population that died from any cause by 1.23 (95% CI: 0.11 to 2.35, p=0.030) percentage points (**Figure 4**), which corresponds to a relative reduction of 2.4%. As for deaths due to dementia, the effect was larger among women than men (**Figure S4**, **Table S3**). Among women, HZ vaccination eligibility reduced all-cause mortality by 1.41 (95% CI: 0.20 to 2.32, p=0.002) percentage points, translating to a relative decrease of 3.0%. The effect was not statistically significant among men (−0.68 [95% CI: −2.37 to 1.01] percentage points, p=0.433).

**Figure 4.**
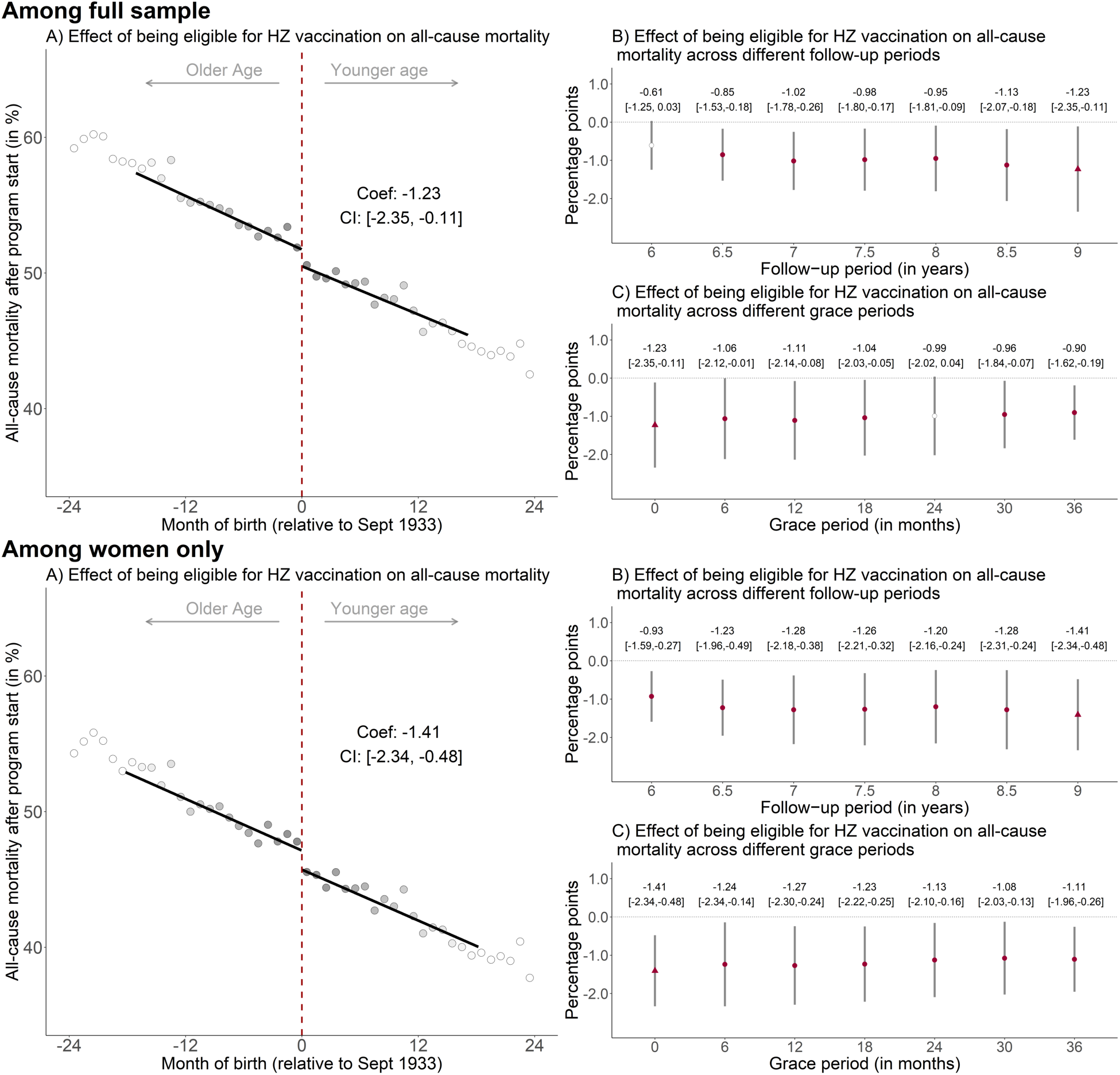
The effect of being eligible for HZ vaccination on the percent of the population alive on September 1 2013 that died from any cause, among the full sample and among women only.^1,2,3,4,5,6,7^. ^1^ Triangles (rather than points) depict our primary specification. ^2^ Red (as opposed to white) fillings denote statistical significance (p<0.05). ^3^ The grey shading of the dots is in proportion to the weight that observations from this birth-month received in the analysis. ^4^ With “grace periods” we refer to time periods since September 1 2013 after which follow-up time is considered to begin to allow for the time needed for HZ vaccination eligibility to begin affecting the outcome. ^5^ Grey vertical bars depict 95% confidence intervals. ^6^ In Panel A, the sample size in the mean squared error-optimal bandwidth is 933,692 and 550,680 for the analysis in the full sample and among women only, respectively. ^7^ The linear regression lines are drawn only in the mean squared error-optimal bandwidth. Abbreviations: Coef=coefficient; CI=95% confidence interval; HZ=herpes zoster; Sept=September.

### Attributing effects on all-cause mortality to effects on deaths due to dementia

There was no significant effect from being eligible for HZ vaccination on deaths that did not have dementia as their underlying cause (−0.72 [95% CI: −1.61 to 0.15] percentage points, p=0.108, **Table S2**). When analyzing deaths that did neither have dementia nor cerebrovascular disease as their underlying cause, the point estimate shifted further towards zero (−0.52 [95% CI: −1.49 to 0.46] percentage points, p=0.305). Using a Cox regression approach, the standardized absolute risk difference for the effect of being eligible for HZ vaccination on deaths due to dementia over our nine-year follow-up period was −0.28 (95% CI: −0.49 to −0.07, p=0.010) percentage points among the whole population, −0.67 (95% CI: −0.98 to −0.35, p<0.001) percentage points among women, and −0.02 (95% CI: −0.27 to 0.23, p=0.865) percentage points among men (**Table S4**).

### Tests for confounding

Except for deaths due to cerebrovascular disease, we did not identify significant effects from being eligible for HZ vaccination on each of the ten (other than dementia) leading underlying causes of death in our data (**Figure 5**, **Figure S5**, and **Figure S6**). The effect on deaths due to cerebrovascular disease was significant among the whole population (−0.28 [95% CI: −0.52 to −0.05] percentage points, p=0.016) and among men (−0.44 [95% CI: −0.72 to −0.15] percentage points, p=0.002), but not among women (−0.15 [95% CI: −0.41 to 0.10] percentage points, p=0.240).

**Figure 5.**
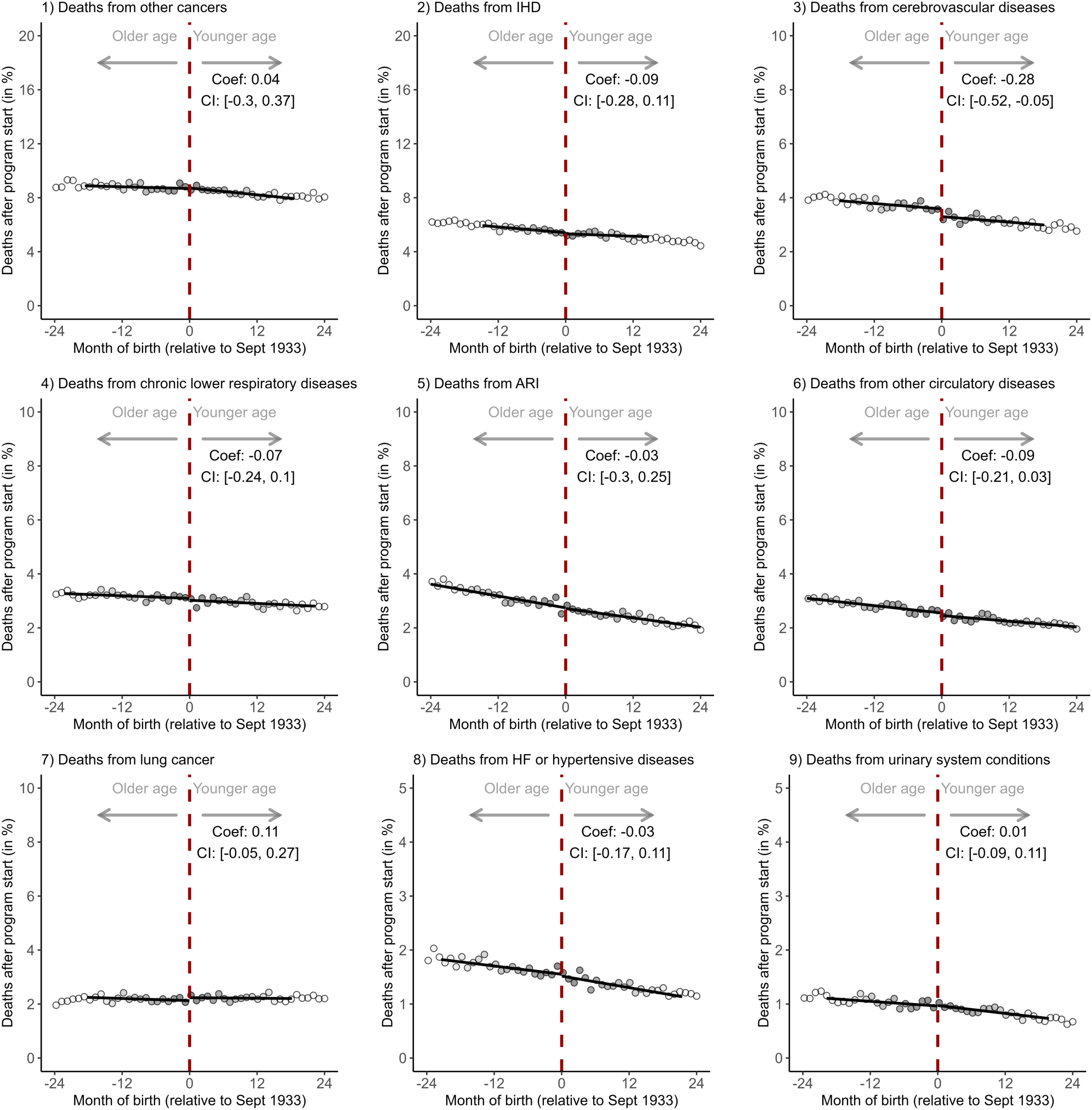
Effects of being eligible for HZ vaccination on the percent of the population alive on September 1 2013 that died from each of the ten (other than dementia) leading underlying causes of death in England and Wales.^1,2,3,4,5,6,7,8^. ^1^ The follow-up period is September 1 2013 to August 31 2022. ^2^ The grey shading of the dots is in proportion to the weight that observations from this birth-month received in the analysis. ^3^ The linear regression lines are drawn only in the mean squared error-optimal bandwidth. ^4^ Other cancers were defined as cancers other than cancers of the trachea, bronchus, and lung. ^5^ Lung cancer encompassed cancers of the trachea, bronchus, and lung. ^6^ Other circulatory diseases were defined as ICD-10 codes I05–I09, I26–I28, I34–I38, I42, I46, I47–I49, I70, and I71. ^7^ Acute respiratory infections were defined as ICD-10 codes J00–J06, J09–J18, and J20–J22. ^8^ The ICD-10 codes to define each cause of death are shown in Table S1. Abbreviations: Coef=coefficient; CI=95% confidence interval; IHD=ischemic heart diseases; ARI=acute respiratory infections; HF=heart failure.

The September 2 date-of-birth threshold was not associated with the proportion of the population that died due to dementia over a nine-year follow-up period in any of the six years preceding the start of the HZ vaccination program (i.e., date-of-birth eligibility thresholds of September 2 for the years 1927 to 1932). This was the case both when examining the whole population (**Figure S7**), as well as when conducting the analyses separately for women and men (**Figure S8**). Similarly, the September 2 date-of-birth threshold was not associated with all-cause mortality in any of the six years preceding the start of the HZ vaccination program, neither for the whole population (**Figure S9**), nor when examining women and men separately (**Figure S10**).

Lastly, there was no significant effect of being eligible for HZ vaccination on the proportion of the population alive on September 1 2004 that died due to dementia in the nine years preceding the start of the HZ vaccination program; neither among the whole population (**Figure S11**), nor among women and men (**Figure S12**). This was also the case for all-cause mortality (**Figure S13** and **S14**).

### Robustness checks

The effect of being eligible for HZ vaccination on reducing the proportion of the population that died due to dementia was somewhat larger when using local squared instead of local linear regression (−0.58 [95% CI: −0.99 to −0.17] percentage points, p=0.006 versus −0.38 [95% CI: −0.68 to −0.08] percentage points, p=0.012; **Figure S15**). When using local squared regression, the effect on all-cause mortality became non-significant among the full sample (**Figure S16**), but remained significant among women (**Figure S17**). Our point estimates for the effect of being eligible for HZ vaccination on the proportion of the population alive on September 1 2013 that died due to dementia remained similar when using different bandwidths (**Figure S18**). This also held true for all-cause mortality (**Figure S19**). Figures S18 and S19 additionally display these estimates in three-month increments for each date-of-birth threshold ranging from September 2 1930 to September 2 1936, showing that the only threshold with significant effects is the threshold that was used for HZ vaccination eligibility.

### Additional analyses

Our negative control outcome analyses found that HZ vaccination eligibility significantly reduced deaths due to cerebrovascular disease for the whole population and among men only. Consistent with the hypothesis that this effect was driven by an aversion of cerebrovascular disease deaths that occurred as a short- to medium-term complication of a shingles episode, the relative effect sizes for this effect decreased over the duration of the follow-up period (**Figure S20**). However, this trend was less pronounced among men (**Figure S21**). The effect estimates for cerebrovascular disease deaths using local squared instead of local linear regression and across different bandwidth choices are shown in **Figures S22, S23,** and **S24**.

## Discussion

We estimated that being eligible for HZ vaccination averted approximately one in twenty deaths that had dementia recorded as their underlying cause over a nine-year follow-up period. Under the assumption that the abrupt increase in HZ vaccination receipt at the September 2 1933 date-of-birth eligibility threshold was similar in magnitude in England as we have reported in country-wide electronic health record data from Wales(13), the effect of actually receiving HZ vaccination (as opposed to merely being eligible) would be an approximately ten percent relative reduction in deaths due to dementia over our nine-year follow-up period. As in our prior study(13), the effect was stronger among women than men. This effect heterogeneity by gender is unlikely explained by differences in HZ vaccination uptake given that our analysis in Welsh electronic health record data found a similar change in HZ vaccination uptake at the September 2 1933 eligibility threshold between women and men(13). Instead, we hypothesize that it could arise from immunological sex differences, given that shingles is more common among women than men(36,37), off-target effects of vaccines have consistently been found to be far stronger among female than male individuals(11), and the immunological differences by sex in the response to vaccines more generally(41).

We found that being eligible for HZ vaccination also reduced all-cause mortality. The main difficulty in attributing this reduction in all-cause mortality to an effect on the dementia disease process is the underestimation of deaths attributable to dementia when using death certificate data on primary causes of death(32). Thus, even if HZ vaccination eligibility had solely reduced deaths caused by dementia, we would still expect some decrease in those deaths not recorded as having dementia as an underlying cause. Nonetheless, deaths due to dementia averted through HZ vaccination eligibility appear to generally not have been replaced by deaths from other causes. If eligibility for HZ vaccination had merely reduced the frequency of mentions of dementia as the underlying cause on death certificates without increasing remaining life expectancy, then we should have found that HZ vaccination eligibility had no effect on all-cause mortality and increased deaths that did not have dementia as their underlying cause. Instead, we found a significant decrease in all-cause mortality from HZ vaccination eligibility and no effect on non-dementia deaths. Our Cox regression approach found similar effect magnitudes for reducing deaths due to dementia as our regression discontinuity approach (−0.28 versus −0.38 percentage points among the whole population, −0.67 versus −0.62 percentage points among women, and −0.02 versus −0.11 percentage points among men). This analysis, which more formally modelled the competing risk between dementia and non-dementia deaths, thus, also supports the conclusion that our observed all-cause mortality effects can generally be attributed to the reduction in deaths due to dementia.

We believe that our estimates are likely to be causal rather than associational for several key reasons. First and foremost, the September 2 1933 eligibility threshold for HZ vaccination is arbitrary such that individuals born shortly before versus shortly after this threshold are in expectation identical to each other in both observed and unobserved potential confounding variables. The use of this “natural randomization” is the key difference between our analysis and the existing body of evidence that has simply correlated vaccine receipt with dementia(14); an approach that is prone to selection bias given the many (and often unmeasured) differences between individuals who are vaccinated and those who are not(15). Second, our month-of-birth cohorts were well balanced in their occurrence of past deaths from any cause, deaths due to dementia, as well as deaths due to other common causes, supporting the assumption that individuals did not differ in their characteristics across the September 2 date-of-birth eligibility threshold. This assumption is further supported by our observations in Welsh electronic health record data that individuals across the September 2 1933 threshold did not differ in their past uptake of preventive health services, incidence of dementia, and diagnoses of the ten leading causes of mortality and disability-adjusted life years(13). Third, it is unlikely that a different relevant intervention or policy used the September 2 1933 threshold as its date-of-birth eligibility criterion because i) there was no difference in deaths due to dementia in the nine-years preceding the HZ vaccination program across the September 2 1933 date-of-birth eligibility threshold; and ii) the September date-of-birth threshold only had an effect on deaths due to dementia in the date-of-birth year of 1933. Lastly, when shifting the date-of-birth eligibility threshold repeatedly by three months ranging from 36 months before to 36 months after September 1933, we only observed significant effects on deaths due to dementia for September 1933.

It is possible that HZ vaccination presented an opportunity for the health system to diagnose dementia. Whether or not an individual had dementia at the time of death may, therefore, be more likely known among the month-of-birth cohorts that were eligible for HZ vaccination. Crucially for the interpretation of our results, this bias would only lead us to under-but not overestimate the beneficial effect of HZ vaccination for preventing deaths due to dementia. It is also possible that the occurrence of shingles presented an opportunity for a diagnosis of dementia. If this was an important source of bias in our analysis, then we would however expect a shingles episode to be an opportunity for chronic disease diagnoses more generally, likely leading us to observe effects of HZ vaccination eligibility not merely on dementia and cerebrovascular disease but also other common causes of death. Further support for the assumption that this was not an important source of bias comes from our previous analysis in Welsh electronic health record data(13), in which there was no effect from HZ vaccination eligibility on chronic disease diagnoses other than dementia and adjustment for the frequency of health service utilization during the follow-up period did not change the effect estimates. Lastly, it is possible that HZ vaccination led to uptake of other preventive tools (e.g., other immunizations or blood pressure medications). In our previous analysis(13), we, however, observed no effect of HZ vaccination on the uptake of influenza immunization, statin use, use of antihypertensive medications, or breast cancer screening participation, nor did we find an effect on health outcomes other than dementia.

Because transient ischemic attacks and strokes are a known complication of shingles (thought to arise from varicella zoster virus-induced vasculopathy affecting the intracerebral arteries(43–45)) and the large number of negative control outcome analyses that we conducted (and, thus, the increased probability of obtaining a significant finding by chance), we believe that our significant findings for cerebrovascular disease deaths are unlikely to be an indication of confounding. It is unclear whether or not fatalities from cerebrovascular disease lie on the causal chain that links HZ vaccination to deaths from dementia. On the one hand, strokes are a well-established risk factor for dementia(48), and a substantial proportion of dementia cases are thought to be due to atherosclerosis of the vasculature(21,49,50), which is also the primary pathophysiological process underlying ischemic strokes(51). On the other hand, the stroke fatality-reducing effects from HZ vaccination eligibility in our analysis were less pronounced and not statistically significant among women, whereas this effect heterogeneity by gender was in the opposite direction for deaths due to dementia. Although the absolute effect magnitudes that we estimated for deaths due to cerebrovascular disease were generally moderate in size (−0.28 percentage points in the whole population), they are nonetheless important from a public health perspective, especially when considering the relative effect sizes (−8.0%) and the subgroup of men (0.44 percentage point absolute and 11.9% relative reduction).

We have previously shown that HZ vaccination reduced the incidence of new dementia diagnoses(13). Because the follow-up period in this prior analysis was only seven years, it was unclear whether HZ vaccination decreased (or delayed) the transition from normal cognitive functioning to dementia, the transition from mild cognitive impairment to dementia, or the deterioration in cognitive function among those who already had mild-to-moderate dementia but were not yet diagnosed. Given the long duration of the natural history of dementia compared to our follow-up period of nine years(16), and the fact that this present study analyzed deaths with dementia as their underlying (rather than contributing) cause of death, our findings in this analysis of mortality data suggest that HZ vaccination had a beneficial effect on cognitive functioning among those who already had dementia. As such, this study suggests that HZ vaccination has (possibly in addition to being a preventive tool) a beneficial effect on cognitive function among individuals who are already at a relatively advanced stage of dementia.

In our view, our repeated finding of the benefits of HZ vaccination for the dementia disease process calls for investments into research to uncover relevant causal mechanisms. Such mechanisms could act through the prevention of clinical and subclinical reactivations of the varicella zoster virus and more indirect immunological pathways, some of which could be specific to live-attenuated vaccines. There is already a considerable body of evidence for these mechanistic hypotheses(6,10), including the finding that herpesviruses seed β-amyloid in mice(7), that reactivations of the varicella zoster virus appear to reactivate the herpes simplex virus-1 in the brain(52), and the evidence from a broad array of research that live-attenuated vaccines induce powerful innate immune mechanisms that have important off-target health effects(10,11). Off-target effects of vaccines have consistently been observed to be far stronger among women than men(10,11), but the mechanisms underlying these sex differences are not well understood and, thus, also represent important lines of future investigation(12).

Our study has several limitations. First, death certificate data is likely to provide an underestimate of deaths caused by dementia. Importantly, however, there is no reason for which the measurement error of deaths due to dementia should differ between those born shortly before versus shortly after the September 2 1933 date-of-birth eligibility threshold. As such, our relative effect estimates are unlikely to be affected by this underreporting. Similarly, potential changes in the accuracy of death certificate data over time are not a source of bias in this study because they affect population groups born on either side of the date-of-birth eligibility threshold for HZ vaccination equally. Second, we did not have information on which individuals received HZ vaccination. As such, our data only allowed us to determine the effect of being eligible for HZ vaccination on deaths due to dementia, but not the effect of actually receiving the HZ vaccine. Calculating the effect of receiving the HZ vaccine would require us to assume that the magnitude of the abrupt change in HZ vaccination receipt at the September 2 1933 date-of-birth eligibility threshold was similar in England as we have observed in Wales (47.2 percentage points)(13). Despite this lacking information on vaccination receipt, we can be confident that HZ vaccination generally occurred prior to our outcome ascertainment because eligibility for HZ vaccination for individuals born between September 2 1933 and September 1 1934 (the year-of-birth cohort that received the highest weight among eligible cohorts in our analyses) lasted for 12 months only and our effect sizes were consistent across different grace periods. Third, our effect size estimates only apply to the age groups around the September 2 1933 date-of-birth threshold. Fourth, our analyses of the effect of HZ vaccination eligibility on deaths due to specific types of dementia were severely limited by the reduced statistical power to detect effects on these rarer outcomes, the neuropathological overlap between dementia types(21–23), and the difficulty of distinguishing dementia types clinically. Lastly, because the new recombinant subunit HZ vaccine (“Shingrix”) was only introduced in the UK in late 2021(53), our findings apply to Zostavax only.

In conclusion, taking advantage of a unique natural randomization in the UK, this study provides evidence that HZ vaccination has a beneficial effect on cognitive function among individuals with dementia. In ascertaining the effect of HZ vaccination eligibility using a different type of data, study population, and outcome, this analysis also represents an important confirmation of our previous finding in Welsh electronic health record data that HZ vaccination has a beneficial effect on the dementia disease process more generally.

## Ethics

This analysis was approved and considered minimal risk by the Stanford University Institutional Review Board on June 9 2023 (protocol number: 70277).

## Acknowledgements

We would like to thank the UK’s Office for National Statistics for providing us with the data used in this study.

## Authors’ contributions

F.M. devised the methodology, analyzed and processed the data, created data visualizations, interpreted the results, and reviewed and edited the original draft. M.X. devised the methodology, interpreted the results, and reviewed and edited the original draft. M.E. devised the methodology, interpreted the results, and reviewed and edited the original draft. S.H. devised the methodology, interpreted the results, and reviewed and edited the original draft. S.C. analyzed and processed the data, created data visualizations, and interpreted the results. P.G. conceived the study, acquired funding, devised the methodology, was responsible for administration and supervision, interpreted the results, and wrote the original draft.

## Data and materials availability

All data and statistical analysis code (in R) will be made available in a publicly accessible GitHub repository upon acceptance of the manuscript for publication.

