## Supplementary Material for "The effect of herpes zoster vaccination on the occurrence of deaths due to dementia in England and Wales"

**Text S1. Description of the herpes zoster vaccination rollout in the United Kingdom.**

The Zostavax vaccine was made available to eligible individuals in England and Wales through a staggered rollout system starting on September 1 2013. Individuals aged 71 years or older were categorized into three groups on September 1 of each year:

i) an ineligible cohort of those aged 71 to 78 years (or 77 years, depending on the year of the program), who will become eligible in the future,
ii) a catch-up cohort, consisting of individuals aged 79 years (or 78 years, again depending on the year of the program),
iii) those who were ineligible as they were aged 80 years or older and who would never again become eligible.

Our study population consisted of adults born between September 1 1925 (88 years old when the program started), and September 1 1941 (72 years old when the program began). Among this group, those born between September 1 1925 and September 1 1933 never became eligible for the vaccine. However, individuals born between September 2 1933 and September 1 1941 gradually became eligible in a catch-up cohort. Specifically, the vaccine was offered to those born between:

- September 2 1933 and September 1 1934 in the first year of the program (September 1 2013 to August 31 2014),
- September 2 1934 and September 1 1936 in the second year (September 1 2014 to August 31 2015),
- September 2 1936 and September 1 1937 in the third year (September 1 2015 to August 31 2016),
- September 2 1937 and September 1 1938 in the fourth year (September 1 2016 to August 31 2017).

Starting from April 1 2017, individuals became eligible for the vaccine on their 78th birthday and remained eligible until their 80th birthday.

**Text S2. Description of regression models.**

Main regression discontinuity model:

$$Y_t = \alpha + \beta_1 \cdot (MOB_t - c_0) + \beta_2 D_t + \beta_3 D_t \cdot (MOB_t - c_0) + \epsilon_t$$

where t indexes the month of birth and s indexes the gender. Y is the number of deaths during the follow-up time divided by the size of the population at the beginning of the follow-up time. The binary variable D indicates eligibility for herpes zoster vaccination (it is, thus, equal to one after the cutoff date of September 2 1933). The term (MOB-$c_0$) denotes the month of birth, centered around the September 2 1933 eligibility date. The interaction term D*(MOB-$c_0$) allows for the slope of the regression line to differ on either side of the date-of-birth eligibility threshold. The parameter $\beta_2$ identifies the effect of herpes zoster vaccination eligibility on the outcome.

Effect heterogeneity by gender:

$$Y_{ts} = \alpha + \beta_1 \cdot (MOB_{ts} - c_0) + \beta_2 D_{ts} + \beta_3 MALE_{ts} + \beta_4 D_{ts} \cdot (MOB_{ts} - c_0) + \beta_5 D_{ts} \cdot MALE_{ts} +$$

$$\beta_6 \cdot (MOB_{ts} - c_0) \cdot MALE_{ts} + \beta_7 D_{ts} \cdot (MOB_{ts} - c_0) \cdot MALE_{ts} + \epsilon_{ts}$$

where t indexes the month of birth and s indexes the gender. Y is the number of deaths during the follow-up time divided by the size of the population at the beginning of the follow-up time. The binary variable D indicates eligibility for herpes zoster vaccination (it is, thus, equal to one after the cutoff date of September 2 1933). The binary variable MALE is equal to one for male month-of-birth observations. The term (MOB-$c_0$) denotes the month of birth, centered around the September 2 1933 eligibility date. The interaction term D*(MOB-$c_0$) allows for the slope of the regression line to differ on either side of the date-of-birth eligibility threshold. Adding the terms (MOB-$c_0$)*MALE and D*(MOB-$c_0$)*MALE allows the slopes to vary by gender. The parameter $\beta_5$ identifies the gender difference in the effect of eligibility for herpes zoster vaccination on the outcome.

Competing risk survival model:

We denote by T the time between a baseline date and the date of an event, and by $E \in (1,2)$ the cause of the event. Let $(E = 1)$ be the event of interest and $(E = 2)$ the competing risk. $(MOB - c_0)$, $D$ and $(MOB - c_0) * D$ are our baseline covariates, where $(MOB - c_0)$ denotes the month of birth, centered around the September 2 1933 eligibility date, and D indicates eligibility for herpes zoster vaccination (it is, thus, equal to one after the cutoff date of September 2 1933).

We consider a setting in which the event time T is right-censored at a fixed time C. We assume that C is conditionally independent of T given ((MOB-$c_0$), D and (MOB-$c_0$)*D) and fix a time $\tau$ such that with near certainty $P(C > \tau | (MOB - c_0), D, (MOB - c_0) * D) > 0$. We denote $\widetilde{\mathrm{T}} = min(T, C)$, $\widetilde{\mathrm{E}} = \Delta E$, and $\Delta = 1(T \leq C)$. $T$ and $E$ are unobserved, and we use $\widetilde{\mathrm{T}}$ and $\widetilde{\mathrm{E}}$ to estimate our model.

Let $S_0(t | (MOB - c_0), D, (MOB - c_0) * D) = P(T > t | (MOB - c_0), D, (MOB - c_0) * D)$ denote the event-free survival function and

$F_j(t | (MOB - c_0), D, (MOB - c_0) * D) = P(T \leq t, E = j | (MOB - c_0), D, (MOB - c_0) * D)$ the cumulative incidence function for event j.

The cause-specific hazard rates are defined as $\lambda_j(t | (MOB - c_0), D, (MOB - c_0) * D) = dF_j(t | (MOB - c_0), D, (MOB - c_0) * D) / S_0(t | (MOB - c_0), D, (MOB - c_0) * D)$.

We also denote the cumulative hazard rates by $\Lambda_j(t|(MOB - c_0), D, (MOB - c_0) * D) = \int_0^t \lambda_j(s|(MOB - c_0), D, (MOB - c_0) * D)ds.$

The stratified Cox regression model (Cox, 1972) for cause j is given by $\lambda_j(t|(MOB - c_0), D, (MOB - c_0) * D) = \lambda_{0j}(t)exp\big(\beta_1(MOB - c_0) + \beta_2 D + \beta_3 D * (MOB - c_0)\big),$

where the betas are log-hazard ratios and $\lambda_{0j}(t)$ is an unspecified baseline hazard function.

We are interested in the cumulative incidence function $F_1$ whose values are the time-dependent absolute probabilities of occurrence of event 1:

$$F_1(t|(MOB - c_0), D, (MOB - c_0) * D) \\ = \int_0^t S(s - |(MOB - c_0), D, (MOB - c_0) * D_t)\lambda_1(s|(MOB - c_0), D, (MOB - c_0) * D)ds$$

where $s -$ denotes the right-sided limit.

We estimate the average treatment effect of HZ vaccine eligibility as

$$E[F_1(t|D = 1, (MOB - c_0), (MOB - c_0) * D) - F_1(t|D = 0, (MOB - c_0), (MOB - c_0) * D)]$$

using the G-formula.

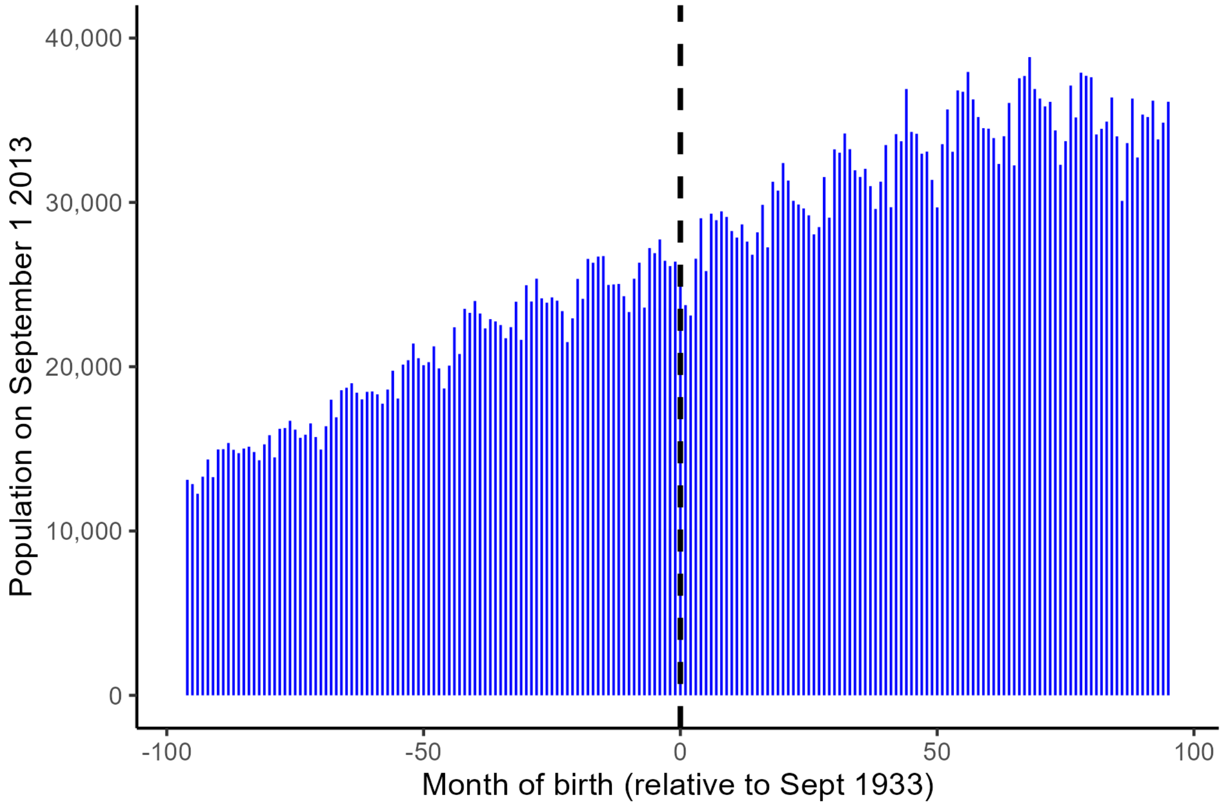


**Figure S1**. Histogram showing the number of individuals in our study population by month of birth.

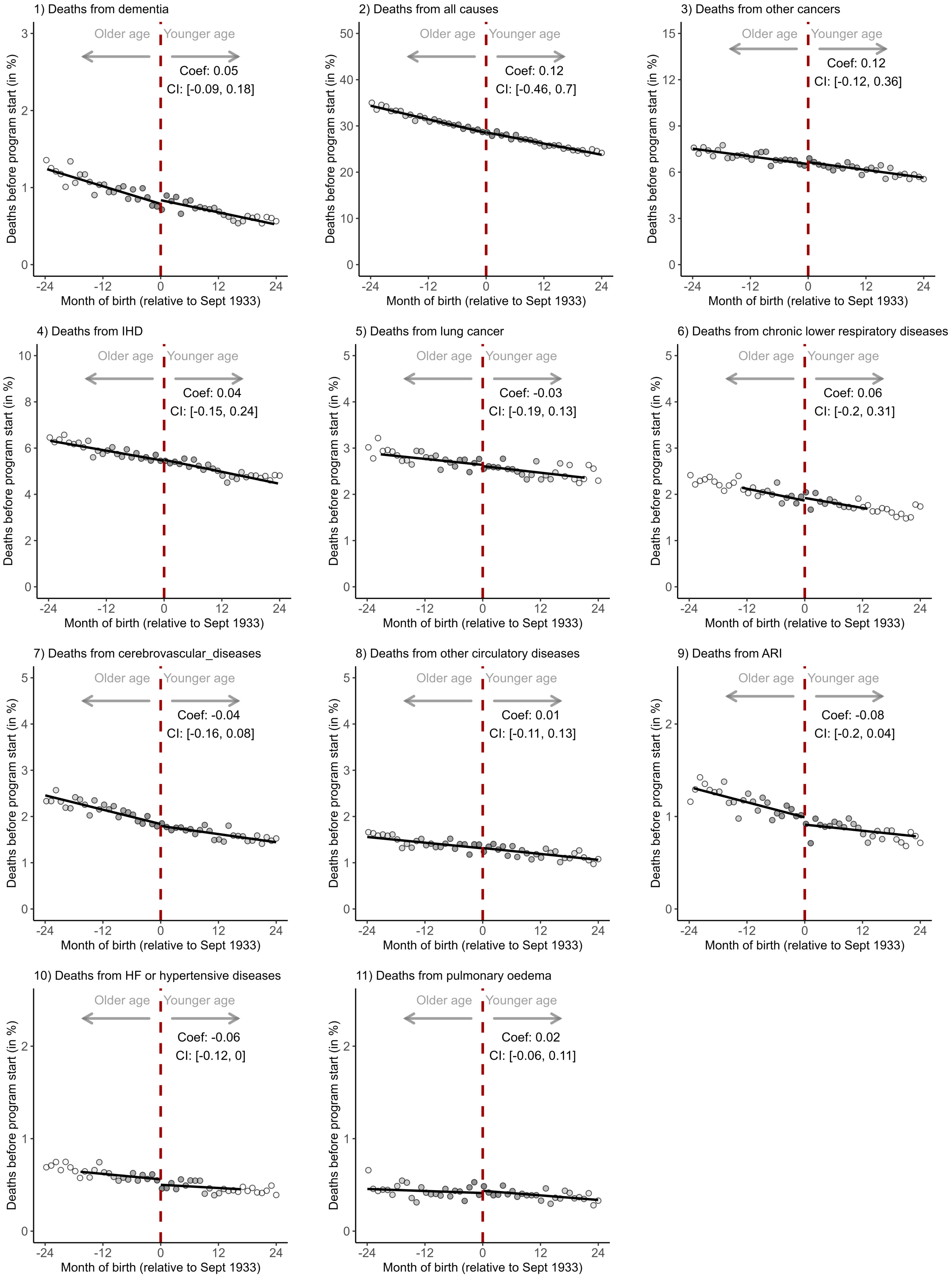


**Figure S2**. Balance tests across the date-of-birth eligibility threshold for herpes zoster vaccination among men.[1,2,3,4,5,6,7,8]

[1] The y-axis shows the percent of the population in each month-of-birth cohort alive on September 1 2004 that died from the given cause in the nine years preceding the start of the herpes zoster vaccination program.
[2] The grey shading of the dots is in proportion to the weight that observations from this birth-month received in the analysis.
[3] The linear regression lines are drawn only in the mean-squared-error optimal bandwidth.
[4] Other cancers were defined as cancers other than cancers of the trachea, bronchus, and lung.
[5] Lung cancer encompassed cancers of the trachea, bronchus, and lung.
[6] Other circulatory diseases were defined as ICD-10 codes I05–I09, I26–I28, I34–I38, I42, I46, I47–I49, I70, and I71.
[7] Acute respiratory infections were defined as ICD-10 codes J00–J06, J09–J18, and J20–J22.
[8] The ICD-10 codes to define each cause of death are shown in Table S1.
Abbreviations: Coef=coefficient; CI=95% confidence interval; IHD=ischemic heart diseases; ARI=acute respiratory infections; HF=heart failure

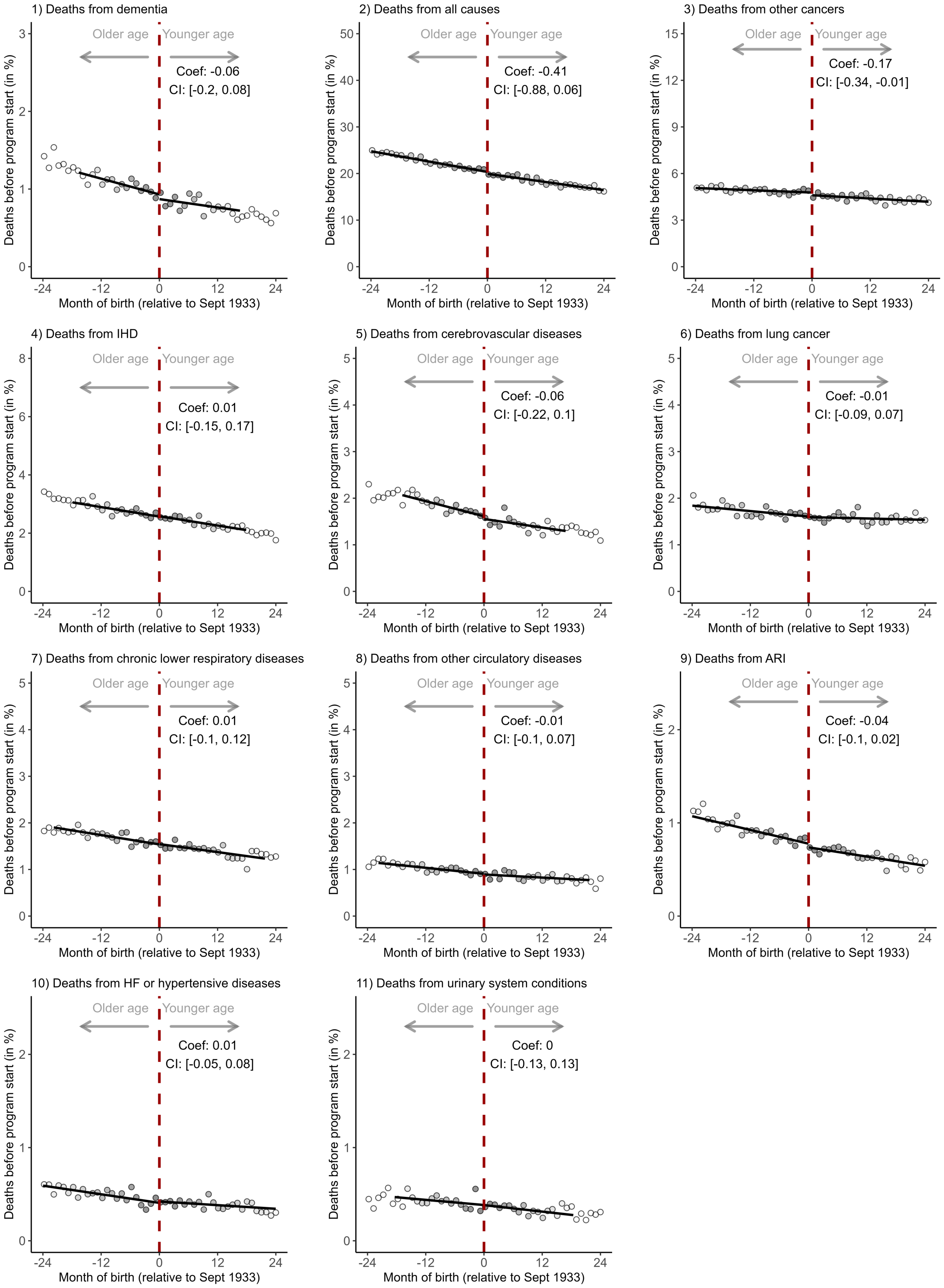


**Figure S3**. Balance tests across the date-of-birth eligibility threshold for herpes zoster vaccination among women.[1,2,3,4,5,6,7,8]

[1] The y-axis shows the percent of the population in each month-of-birth cohort alive on September 1 2004 that died from the given cause in the nine years preceding the start of the herpes zoster vaccination program.
[2] The grey shading of the dots is in proportion to the weight that observations from this birth-month received in the analysis.
[3] The linear regression lines are drawn only in the mean-squared-error optimal bandwidth.
[4] Other cancers were defined as cancers other than cancers of the trachea, bronchus, and lung.
[5] Lung cancer encompassed cancers of the trachea, bronchus, and lung.
[6] Other circulatory diseases were defined as ICD-10 codes I05–I09, I26–I28, I34–I38, I42, I46, I47–I49, I70, and I71.
[7] Acute respiratory infections were defined as ICD-10 codes J00–J06, J09–J18, and J20–J22.
[8] The ICD-10 codes to define each cause of death are shown in Table S1.
Abbreviations: Coef=coefficient; CI=95% confidence interval; IHD=ischemic heart diseases; ARI=acute respiratory infections; HF=heart failure

### Men

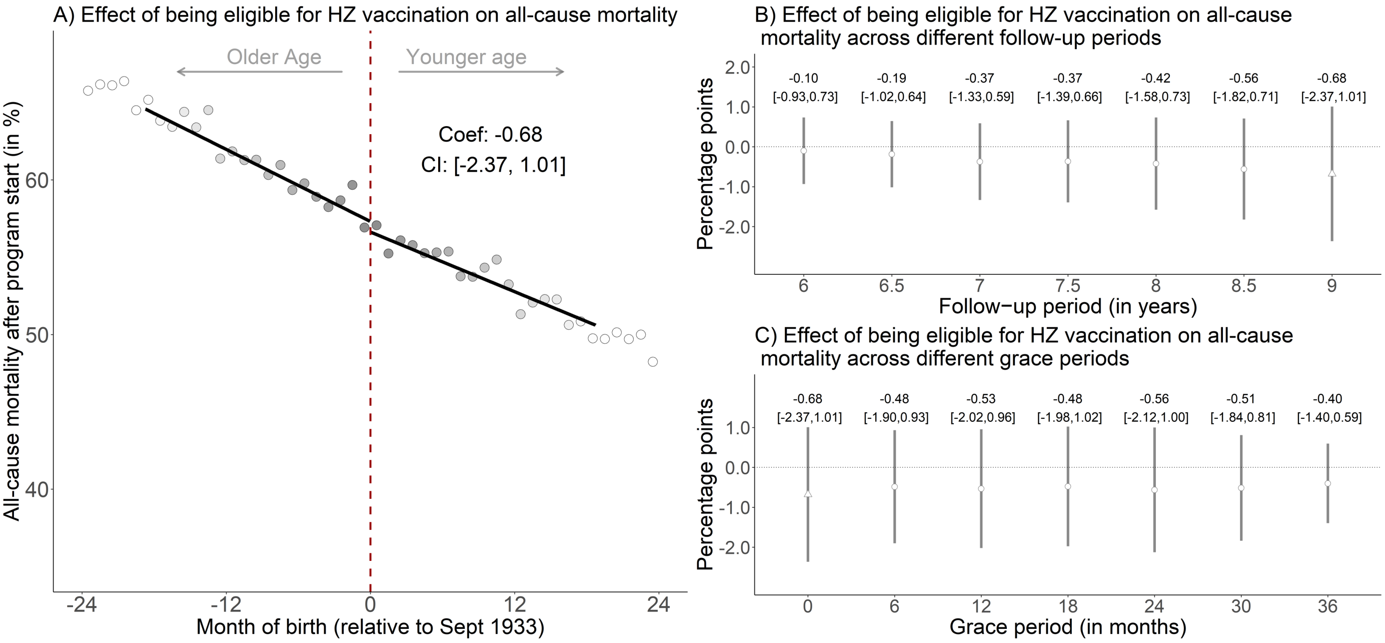


**Figure S4**. The effect of being eligible for herpes zoster vaccination on the percent of the population alive on September 1 2013 that died from any cause, among men only.[1,2,3,4,5,6,7]

[1] Triangles (rather than points) depict our primary specification.
[2] Red (as opposed to white) fillings denote statistical significance (p<0.05).
[3] The grey shading of the dots is in proportion to the weight that observations from this birth-month received in the analysis.
[4] With “grace periods” we refer to time periods since September 1 2013 after which follow-up time is considered to begin to allow for the time needed for herpes zoster vaccination eligibility to begin affecting the outcome.
[5] Grey vertical bars depict 95% confidence intervals.
[6] In Panel A, the sample size in the mean squared error-optimal bandwidth is 440,831 men.
[7] The linear regression lines are drawn only in the mean squared error-optimal bandwidth.
Abbreviations: Coef=coefficient; CI=95% confidence interval; HZ=herpes zoster; Sept=September

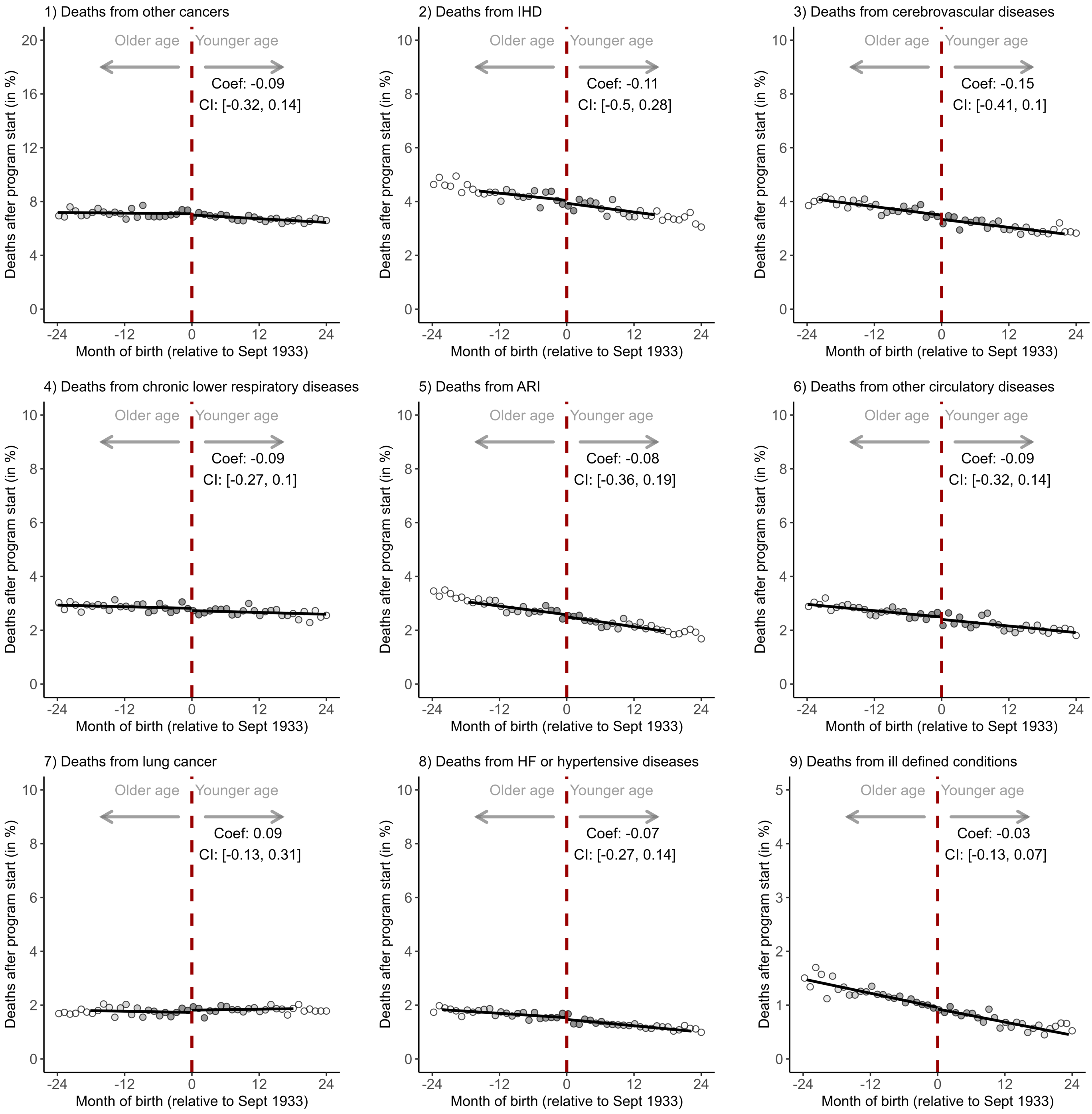


**Figure S5**. Effects among women of being eligible for herpes zoster vaccination on each of the ten (other than dementia) leading underlying causes of death in England and Wales.[1,2,3,4,5,6,7,8]

[1] The follow-up period is September 1 2013 to August 31 2022.
[2] The grey shading of the dots is in proportion to the weight that observations from this birth-month received in the analysis.
[3] The linear regression lines are drawn only in the mean-squared-error optimal bandwidth.
[4] Other cancers were defined as cancers other than cancers of the trachea, bronchus, and lung.
[5] Lung cancer encompassed cancers of the trachea, bronchus, and lung.
[6] Other circulatory diseases were defined as ICD-10 codes I05–I09, I26–I28, I34–I38, I42, I46, I47–I49, I70, and I71.
[7] Acute respiratory infections were defined as ICD-10 codes J00–J06, J09–J18, and J20–J22.
[8] The ICD-10 codes to define each cause of death are shown in Table S1.
Abbreviations: Coef=coefficient; CI=95% confidence interval; IHD=ischemic heart diseases; ARI=acute respiratory infections; HF=heart failure

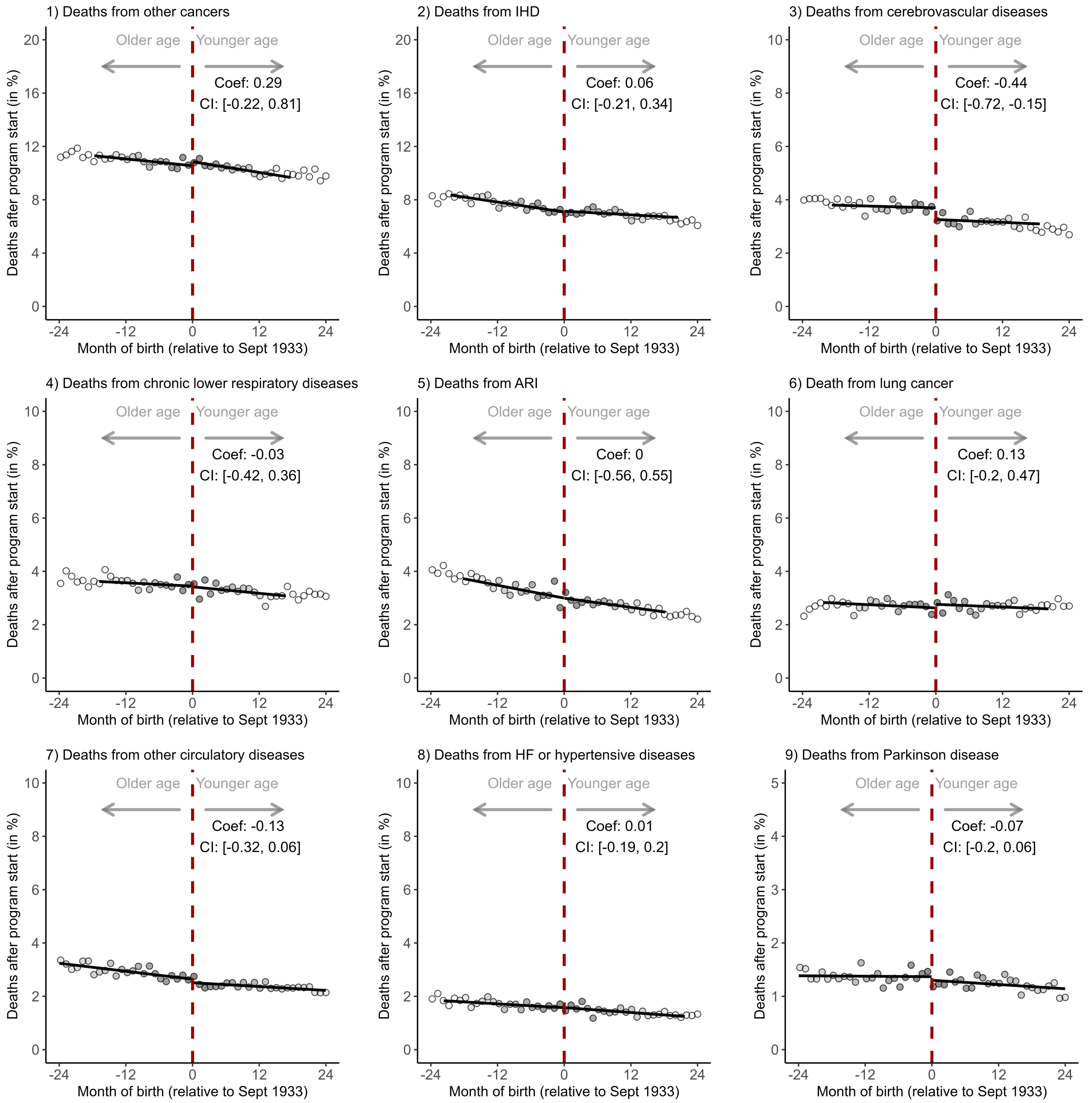


**Figure S6**. Effects among men of being eligible for herpes zoster vaccination on each of the ten (other than dementia) leading underlying causes of death in England and Wales. [1,2,3,4,5,6,7,8]

[1] The follow-up period is September 1 2013 to August 31 2022.
[2] The grey shading of the dots is in proportion to the weight that observations from this birth-month received in the analysis.
[3] The linear regression lines are drawn only in the mean-squared-error optimal bandwidth.
[4] Other cancers were defined as cancers other than cancers of the trachea, bronchus, and lung.
[5] Lung cancer encompassed cancers of the trachea, bronchus, and lung.
[6] Other circulatory diseases were defined as ICD-10 codes I05–I09, I26–I28, I34–I38, I42, I46, I47–I49, I70, and I71.
[7] Acute respiratory infections were defined as ICD-10 codes J00–J06, J09–J18, and J20–J22.
[8] The ICD-10 codes to define each cause of death are shown in Table S1.

Abbreviations: Coef=coefficient; CI=95% confidence interval; IHD=ischemic heart diseases; ARI=acute respiratory infections; HF=heart failure

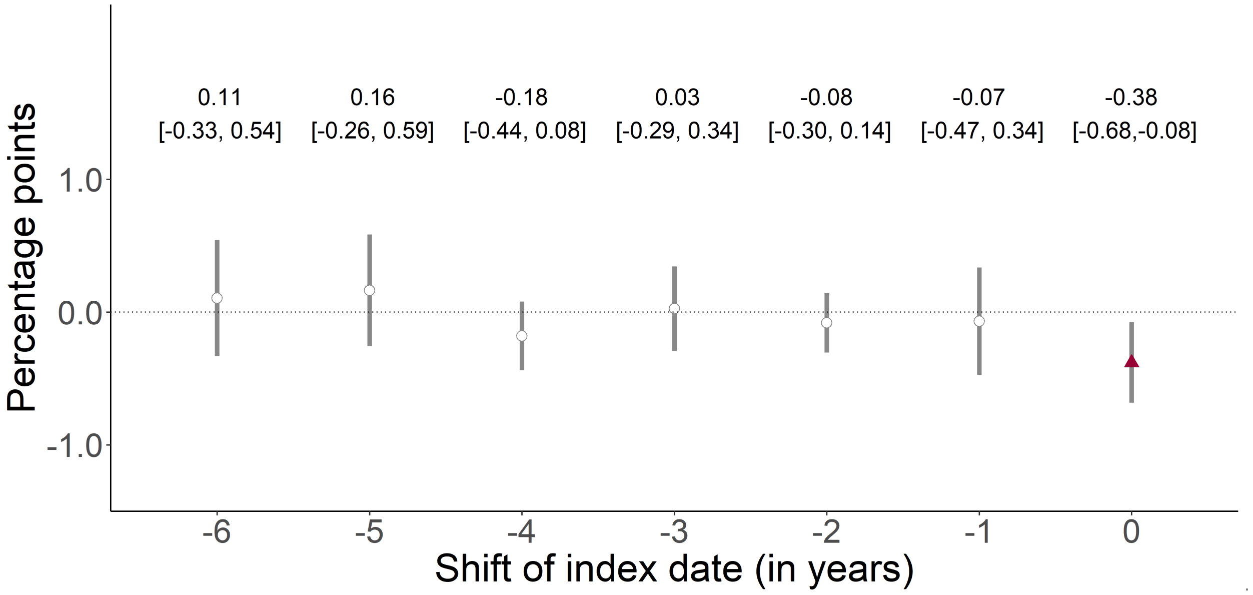


**Figure S7**. The September 2 date-of-birth threshold was not associated with deaths due to dementia over a nine-year follow-up period in any of the six years preceding the start of the herpes zoster vaccination program.[1,2,3,4]

[1] This analysis implemented the identical analysis as for September 1 2013 (the date on which the herpes zoster vaccination program started), for September 1 of each of the six years preceding 2013. For example, when moving the start date of the program to “-2” (i.e., September 1 2011), we started the follow-up period on September 1 2011 and compared individuals around the September 2 1931 eligibility threshold. The purpose of this analysis was to verify that the day-month (i.e., September 2) cutoff used for herpes zoster vaccination eligibility was not also used for other interventions that affected deaths due to dementia.
[2] Red (as opposed to white) fillings denote statistical significance ($p<0.05$).
[3] Grey vertical bars depict 95% confidence intervals.
[4] The number labels state the point estimate and (in square brackets) accompanying 95% confidence interval.

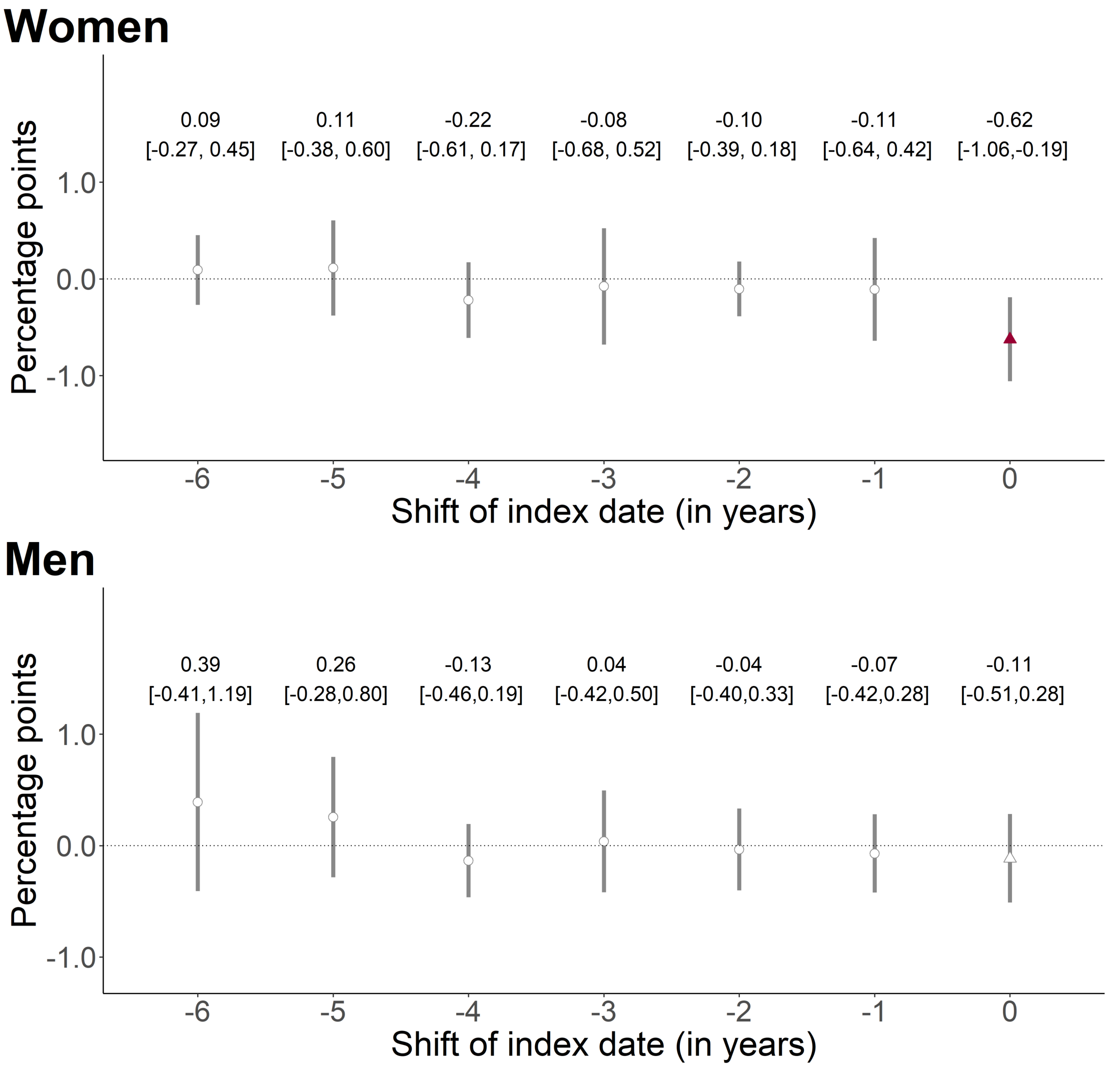


**Figure S8**. The September 2 date-of-birth threshold was not associated with deaths due to dementia over a nine-year follow-up period in any of the six years preceding the start of the herpes zoster vaccination program, separately for women and men.[1,2,3,4]

[1] This analysis implemented the identical analysis as for September 1 2013 (the date on which the herpes zoster vaccination program started), for September 1 of each of the six years preceding 2013. For example, when moving the start date of the program to “-2” (i.e., September 1 2011), we started the follow-up period on September 1 2011 and compared individuals around the September 2 1931 eligibility threshold. The purpose of this analysis was to verify that the day-month (i.e., September 2) cutoff used for herpes zoster vaccination eligibility was not also used for other interventions that affected deaths due to dementia.

[2] Red (as opposed to white) fillings denote statistical significance ($p<0.05$).

[3] Grey vertical bars depict 95% confidence intervals.

[4] The number labels state the point estimate and (in square brackets) accompanying 95% confidence interval.

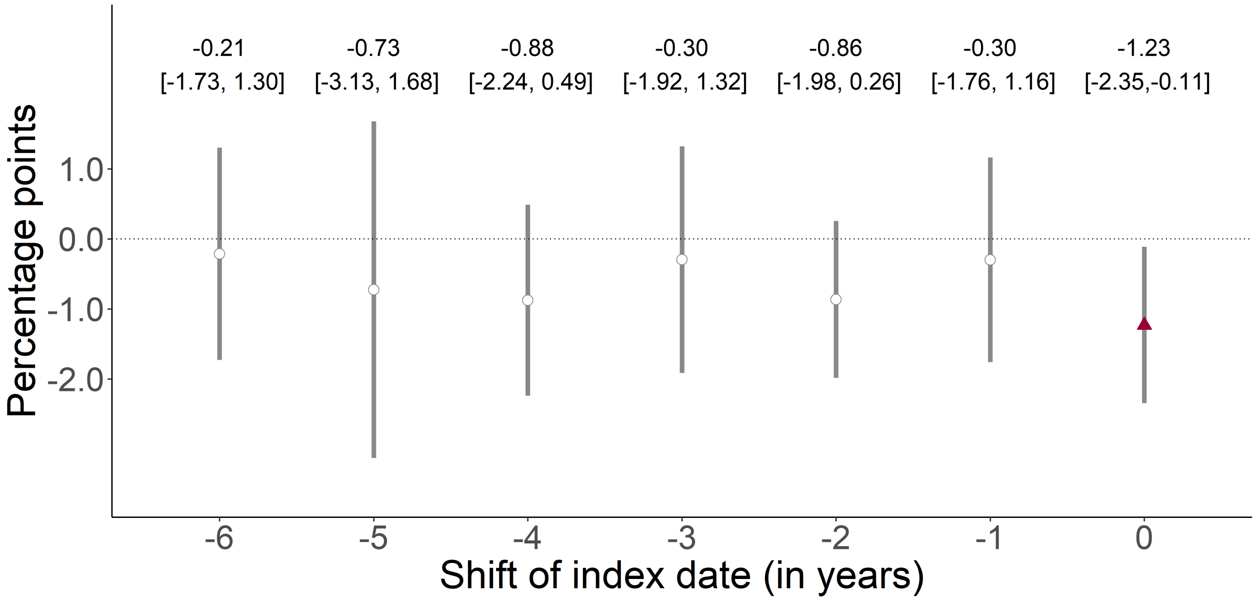


**Figure S9**. The September 2 date-of-birth threshold was not associated with all-cause mortality over a nine-year follow-up period in any of the six years preceding the start of the herpes zoster vaccination program.[1,2,3,4]

[1] This analysis implemented the identical analysis as for September 1 2013 (the date on which the herpes zoster vaccination program started), for September 1 of each of the six years preceding 2013. For example, when moving the start date of the program to “-2” (i.e., September 1 2011), we started the follow-up period on September 1 2011 and compared individuals around the September 2 1931 eligibility threshold. The purpose of this analysis was to verify that the day-month (i.e., September 2) cutoff used for herpes zoster vaccination eligibility was not also used for other interventions that affected all-cause mortality.

[2] Red (as opposed to white) fillings denote statistical significance ($p<0.05$).

[3] Grey vertical bars depict 95% confidence intervals.

[4] The number labels state the point estimate and (in square brackets) accompanying 95% confidence interval.

**Women**

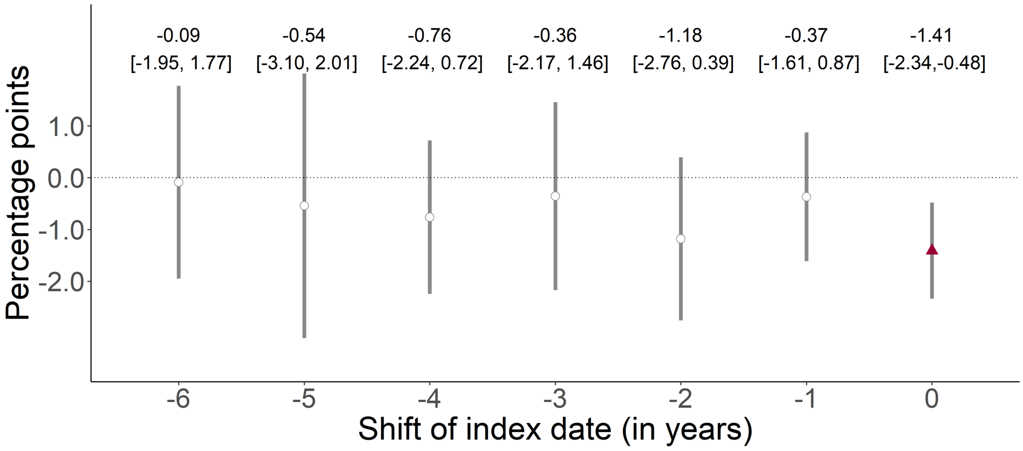


**Men**

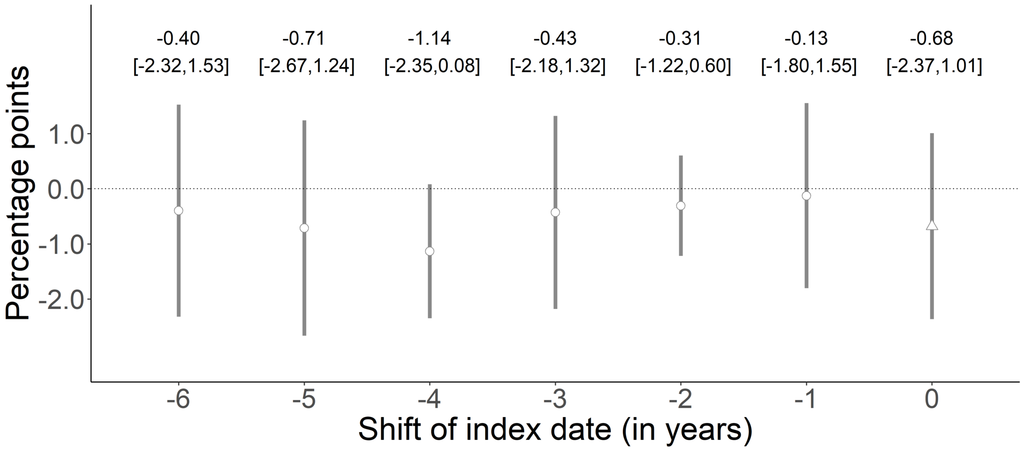


**Figure S10**. The September 2 date-of-birth threshold was not associated with all-cause mortality over a nine-year follow-up period in any of the six years preceding the start of the herpes zoster vaccination program, separately for women and men.[1,2,3,4]

[1] This analysis implemented the identical analysis as for September 1 2013 (the date on which the herpes zoster vaccination program started), for September 1 of each of the six years preceding 2013. For example, when moving the start date of the program to "-2" (i.e., September 1 2011), we started the follow-up period on September 1 2011 and compared individuals around the September 2 1931 eligibility threshold. The purpose of this analysis was to verify that the day-month (i.e., September 2) cutoff used for herpes zoster vaccination eligibility was not also used for other interventions that affected all-cause mortality.

[2] Red (as opposed to white) fillings denote statistical significance ($p<0.05$).

[3] Grey vertical bars depict 95% confidence intervals.

[4] The number labels state the point estimate and (in square brackets) accompanying 95% confidence interval.

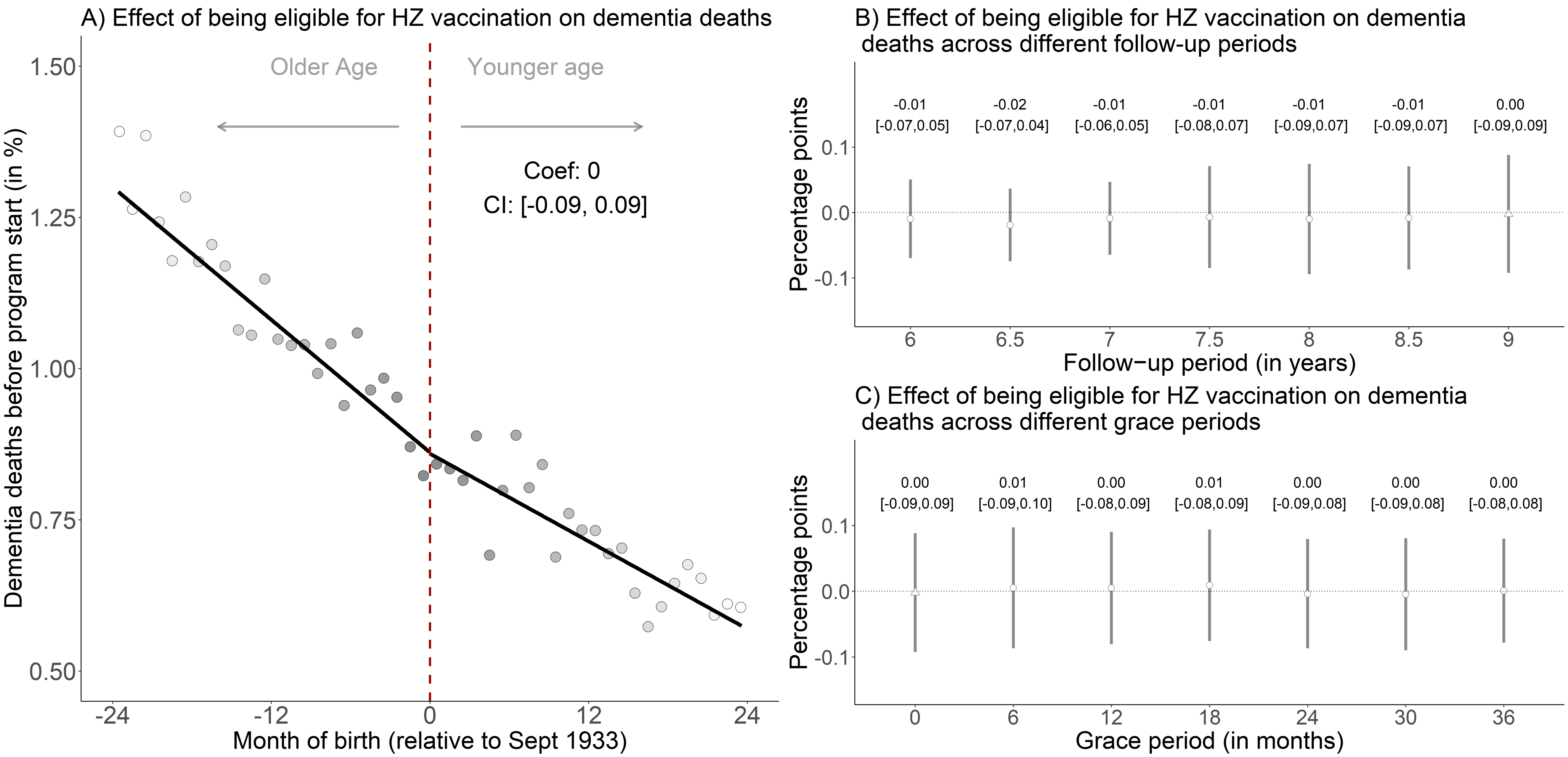


**Figure S11**. No effect on deaths due to dementia in the nine years *prior* to the start of the herpes zoster vaccination program (i.e., between September 1 2004 to August 31 2013).[1,2,3,4,5,6,7,8]

[1] This figure shows the results from the identical analysis as implemented for our primary analysis (for which the results are shown in Fig. 2 in the main manuscript) except that we followed individuals from September 1 2004 to August 31 2013 instead of from September 1 2013 to August 31 2022. This analysis, thus, compared the exact same date-of-birth cohorts to each other and had the same length of follow-up as our primary analysis but used the nine years prior to the start of the herpes zoster vaccine rollout as follow-up period instead of the nine years after program start. The purpose of this analysis was to verify that the same date-of-birth eligibility date used for herpes zoster vaccine eligibility was not used for other interventions in the past that affected dementia risk.

[2] Triangles (rather than points) depict our primary specification.
[3] Red (as opposed to white) fillings denote statistical significance ($p<0.05$).
[4] The grey shading of the dots is in proportion to the weight that observations from this birth-month received in the analysis.
[5] With “grace periods” we refer to time periods since September 1 2004 after which follow-up time is considered to begin.
[6] Grey vertical bars depict 95% confidence intervals.
[7] In Panel A, the sample size in the mean squared error-optimal bandwidth is 1,667,745 adults.
[8] The linear regression lines are drawn only in the mean squared error-optimal bandwidth.
Abbreviations: Coef=coefficient; CI=95% confidence interval; HZ=herpes zoster; Sept=September

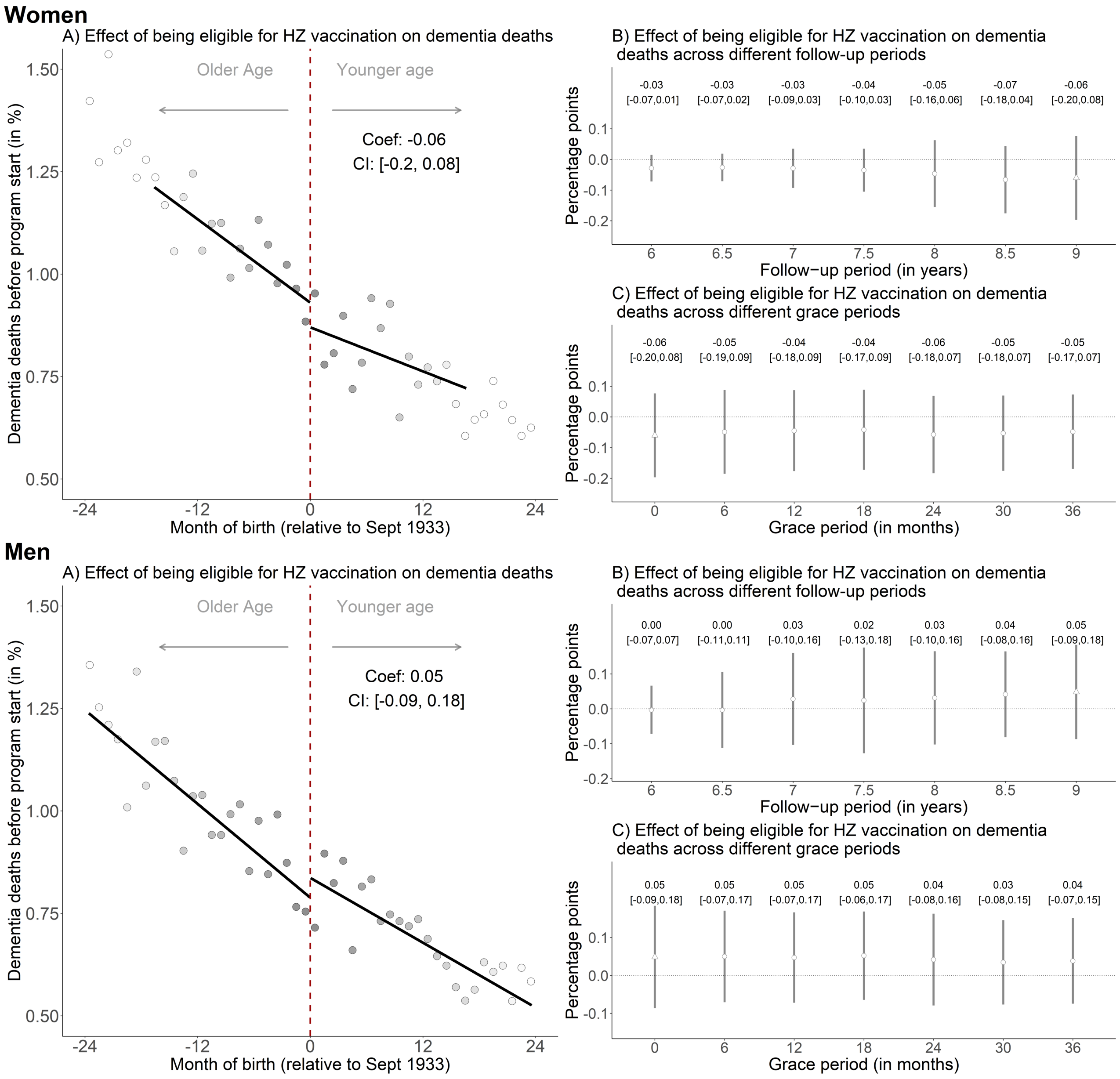
Women
A) Effect of being eligible for HZ vaccination on dementia deaths
Older Age
Younger age
Coef: -0.06
CI: [-0.2, 0.08]
Dementia deaths before program start (in %)
Month of birth (relative to Sept 1933)
B) Effect of being eligible for HZ vaccination on dementia deaths across different follow-up periods
-0.03 [-0.07,0.01]
-0.03 [-0.07,0.02]
-0.03 [-0.09,0.03]
-0.04 [-0.10,0.03]
-0.05 [-0.16,0.06]
-0.07 [-0.18,0.04]
-0.06 [-0.20,0.08]
Percentage points
Follow−up period (in years)
C) Effect of being eligible for HZ vaccination on dementia deaths across different grace periods
-0.06 [-0.20,0.08]
-0.05 [-0.19,0.09]
-0.04 [-0.18,0.09]
-0.04 [-0.17,0.09]
-0.06 [-0.18,0.07]
-0.05 [-0.18,0.07]
-0.05 [-0.17,0.07]
Percentage points
Grace period (in months)
Men
A) Effect of being eligible for HZ vaccination on dementia deaths
Older Age
Younger age
Coef: 0.05
CI: [-0.09, 0.18]
Dementia deaths before program start (in %)
Month of birth (relative to Sept 1933)
B) Effect of being eligible for HZ vaccination on dementia deaths across different follow-up periods
0.00 [-0.07,0.07]
0.00 [-0.11,0.11]
0.03 [-0.10,0.16]
0.02 [-0.13,0.18]
0.03 [-0.10,0.16]
0.04 [-0.08,0.16]
0.05 [-0.09,0.18]
Percentage points
Follow−up period (in years)
C) Effect of being eligible for HZ vaccination on dementia deaths across different grace periods
0.05 [-0.09,0.18]
0.05 [-0.07,0.17]
0.05 [-0.07,0.17]
0.05 [-0.06,0.17]
0.04 [-0.08,0.16]
0.03 [-0.08,0.15]
0.04 [-0.07,0.15]
Percentage points
Grace period (in months)

**Figure S12**. No effect on deaths due to dementia in the nine years *prior* to the start of the herpes zoster vaccination program (i.e., between September 1 2004 to August 31 2013), separately for women and men.[1,2,3,4,5,6,7,8]

[1] This figure shows the results from the identical analysis as implemented for our primary analysis (for which the results are shown in Fig. 2 in the main manuscript) except that we followed individuals from September 1 2004 to August 31 2013 instead of from September 1 2013 to August 31 2022. This analysis, thus, compared the exact same date-of-birth cohorts to each other and had the same length of follow-up as our primary analysis but used the nine years prior to the start of the herpes zoster vaccine rollout as follow-up period instead of the nine years after program start. The purpose of this analysis was to verify that the same date-of-birth eligibility date used for herpes zoster vaccine eligibility was not used for other interventions in the past that affected dementia risk.

[2] Triangles (rather than points) depict our primary specification.

[3] Red (as opposed to white) fillings denote statistical significance ($p<0.05$).

[4] The grey shading of the dots is in proportion to the weight that observations from this birth-month received in the analysis.

[5] With "grace periods" we refer to time periods since September 1 2004 after which follow-up time is considered to begin.

[6] Grey vertical bars depict 95% confidence intervals.

[7] In Panel A, the sample size in the mean squared error-optimal bandwidth is 613,075 and 786,917 for women and men, respectively.

[8] The linear regression lines are drawn only in the mean squared error-optimal bandwidth.

Abbreviations: Coef=coefficient; CI=95% confidence interval; HZ=herpes zoster; Sept=September.

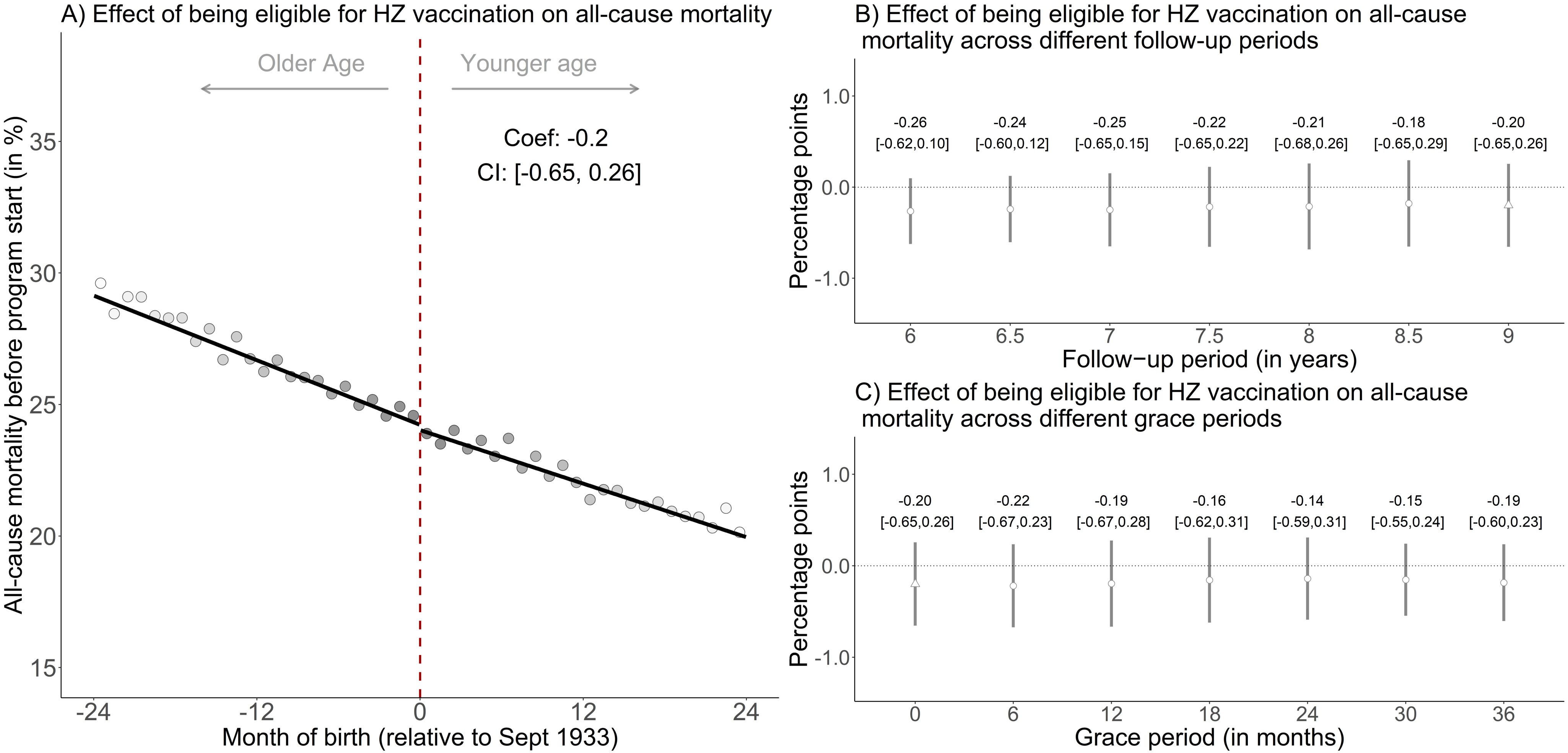


**Figure S13**. No effect on all-cause mortality in the nine years *prior* to the start of the herpes zoster vaccination program (i.e., between September 1 2004 to August 31 2013).[1,2,3,4,5,6,7,8]

[1] This figure shows the results from the identical analysis as implemented for our primary analysis (for which the results are shown in Fig. 4 in the main manuscript) except that we followed individuals from September 1 2004 to August 31 2013 instead of from September 1 2013 to August 31 2022. This analysis thus compared the exact same date-of-birth cohorts to each other and had the same length of follow-up as our primary analysis but used the nine years prior to the start of the herpes zoster vaccine rollout as follow-up period instead of the nine years after program start. The purpose of this analysis was to verify that the same date-of-birth eligibility date used for herpes zoster vaccine eligibility was not used for other interventions in the past that affected mortality risk.

[2] Triangles (rather than points) depict our primary specification.
[3] Red (as opposed to white) fillings denote statistical significance ($p<0.05$).
[4] The grey shading of the dots is in proportion to the weight that observations from this birth-month received in the analysis.
[5] With “grace periods” we refer to time periods since September 1 2013 after which follow-up time is considered to begin to allow for the time needed for herpes zoster vaccination eligibility to begin affecting the outcome.
[6] Grey vertical bars depict 95% confidence intervals.
[7] In Panel A, the sample size in the mean squared error-optimal bandwidth is 1,316,870 adults.
[8] The linear regression lines are drawn only in the mean squared error-optimal bandwidth.
Abbreviations: Coef=coefficient; CI=95% confidence interval; HZ=herpes zoster; Sept=September.

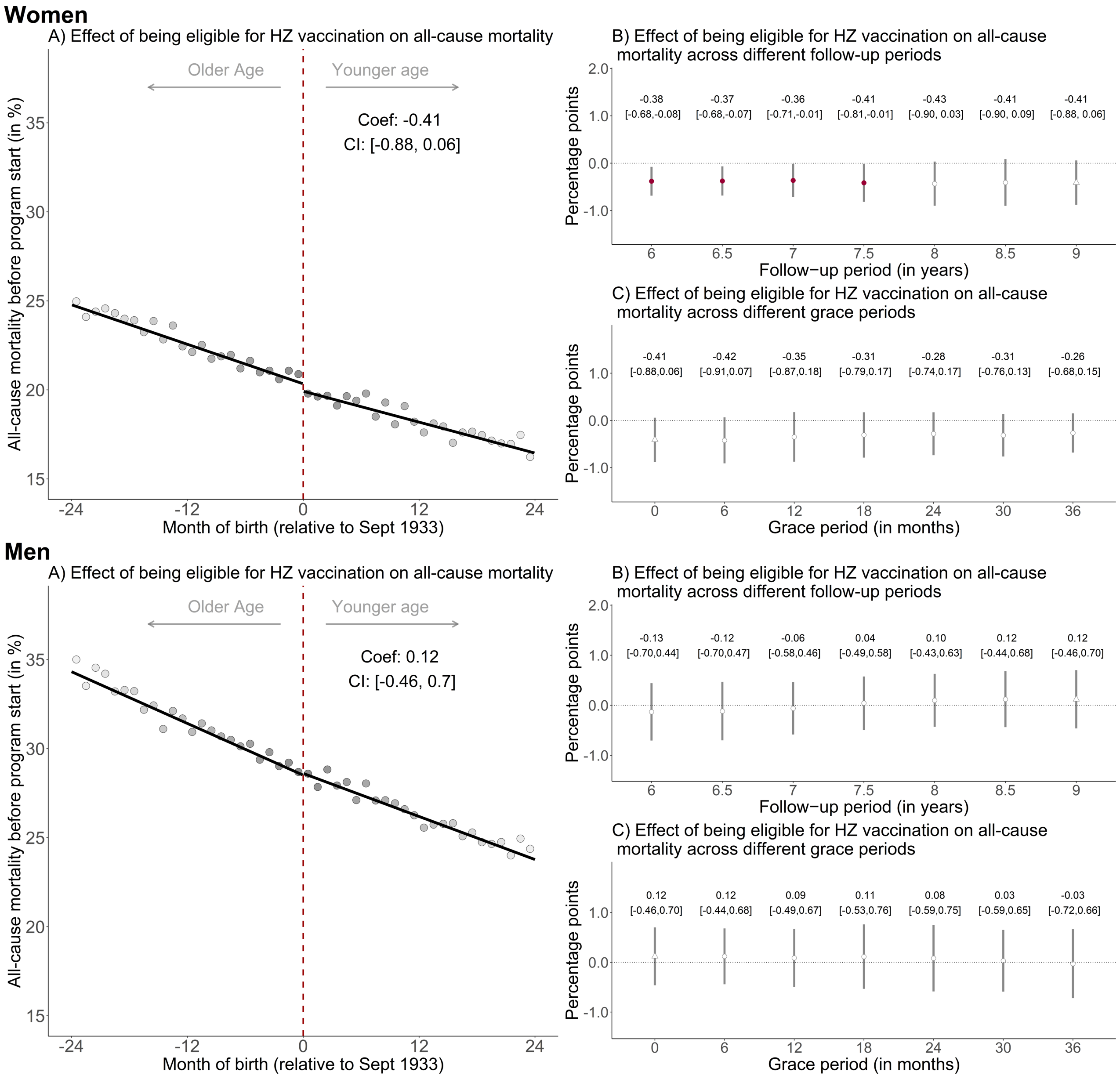
Women
A) Effect of being eligible for HZ vaccination on all-cause mortality
Older Age
Younger age
Coef: -0.41
CI: [-0.88, 0.06]
All-cause mortality before program start (in %)
Month of birth (relative to Sept 1933)
B) Effect of being eligible for HZ vaccination on all-cause mortality across different follow-up periods
-0.38 [-0.68,-0.08]
-0.37 [-0.68,-0.07]
-0.36 [-0.71,-0.01]
-0.41 [-0.81,-0.01]
-0.43 [-0.90, 0.03]
-0.41 [-0.90, 0.09]
-0.41 [-0.88, 0.06]
Percentage points
Follow−up period (in years)
C) Effect of being eligible for HZ vaccination on all-cause mortality across different grace periods
-0.41 [-0.88,0.06]
-0.42 [-0.91,0.07]
-0.35 [-0.87,0.18]
-0.31 [-0.79,0.17]
-0.28 [-0.74,0.17]
-0.31 [-0.76,0.13]
-0.26 [-0.68,0.15]
Percentage points
Grace period (in months)
Men
A) Effect of being eligible for HZ vaccination on all-cause mortality
Older Age
Younger age
Coef: 0.12
CI: [-0.46, 0.7]
All-cause mortality before program start (in %)
Month of birth (relative to Sept 1933)
B) Effect of being eligible for HZ vaccination on all-cause mortality across different follow-up periods
-0.13 [-0.70,0.44]
-0.12 [-0.70,0.47]
-0.06 [-0.58,0.46]
0.04 [-0.49,0.58]
0.10 [-0.43,0.63]
0.12 [-0.44,0.68]
0.12 [-0.46,0.70]
Percentage points
Follow−up period (in years)
C) Effect of being eligible for HZ vaccination on all-cause mortality across different grace periods
0.12 [-0.46,0.70]
0.12 [-0.44,0.68]
0.09 [-0.49,0.67]
0.11 [-0.53,0.76]
0.08 [-0.59,0.75]
0.03 [-0.59,0.65]
-0.03 [-0.72,0.66]
Percentage points
Grace period (in months)

**Figure S14**. No effect on all-cause mortality in the nine years *prior* to the start of the herpes zoster vaccination program (i.e., between September 1 2004 to August 31 2013), separately for women and men.[1,2,3,4,5,6,7,8]

[1] This figure shows the results from the identical analysis as implemented for our primary analysis (for which the results are shown in Fig. 4 in the main manuscript) except that we followed individuals from September 1 2004 to August 31 2013 instead of from September 1 2013 to August 31 2022. This analysis thus compared the exact same date-of-birth cohorts to each other and had the same length of follow-up as our primary analysis but used the nine years prior to the start of the herpes zoster vaccine rollout as follow-up period instead of the nine years after program start. The purpose of this analysis was to verify that the same date-of-birth eligibility date used for herpes zoster vaccine eligibility was not used for other interventions in the past that affected mortality risk.
[2] Triangles (rather than points) depict our primary specification.
[3] Red (as opposed to white) fillings denote statistical significance ($p<0.05$).
[4] The grey shading of the dots is in proportion to the weight that observations from this birth-month received in the analysis.
[5] With “grace periods” we refer to time periods since September 1 2013 after which follow-up time is considered to begin to allow for the time needed for herpes zoster vaccination eligibility to begin affecting the outcome.
[6] Grey vertical bars depict 95% confidence intervals.
[7] In Panel A, the sample size in the mean squared error-optimal bandwidth is 819,679 and 631,842 for women and men, respectively.
[8] The linear regression lines are drawn only in the mean squared error-optimal bandwidth.
Abbreviations: Coef=coefficient; CI=95% confidence interval; HZ=herpes zoster; Sept=September.

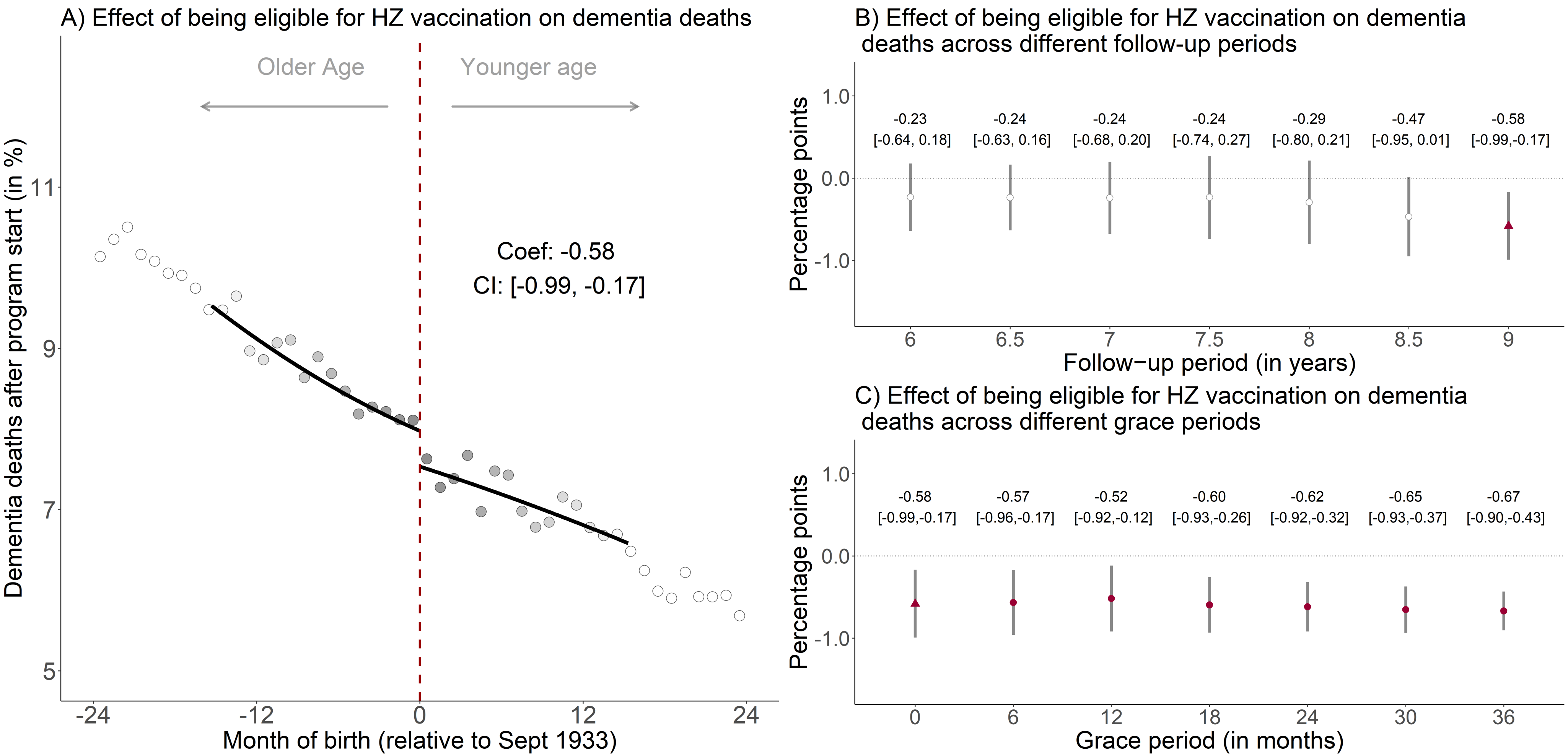


**Figure S15**. The effect of being eligible for herpes zoster vaccination on the percent of the population alive on September 1 2013 that died due to dementia, using local squared instead of local linear regression.[1,2,3,4,5,6,7]

[1] Triangles (rather than points) depict our primary specification.

[2] Red (as opposed to white) fillings denote statistical significance ($p<0.05$).

[3] The grey shading of the dots is in proportion to the weight that observations from this birth-month received in the analysis.

[4] With "grace periods" we refer to time periods since September 1 2013 after which follow-up time is considered to begin to allow for the time needed for herpes zoster vaccination eligibility to begin affecting the outcome.

[5] Grey vertical bars depict 95% confidence intervals, which are estimated following Calonico, Cattaneo, and Titiunik (2014)(1).

[6] In Panel A, the sample size in the mean squared error-optimal bandwidth is 823,548 adults.
[7] The linear regression lines are drawn only in the mean squared error-optimal bandwidth.
Abbreviations: Coef=coefficient; CI=95% confidence interval; HZ=herpes zoster; Sept=September.

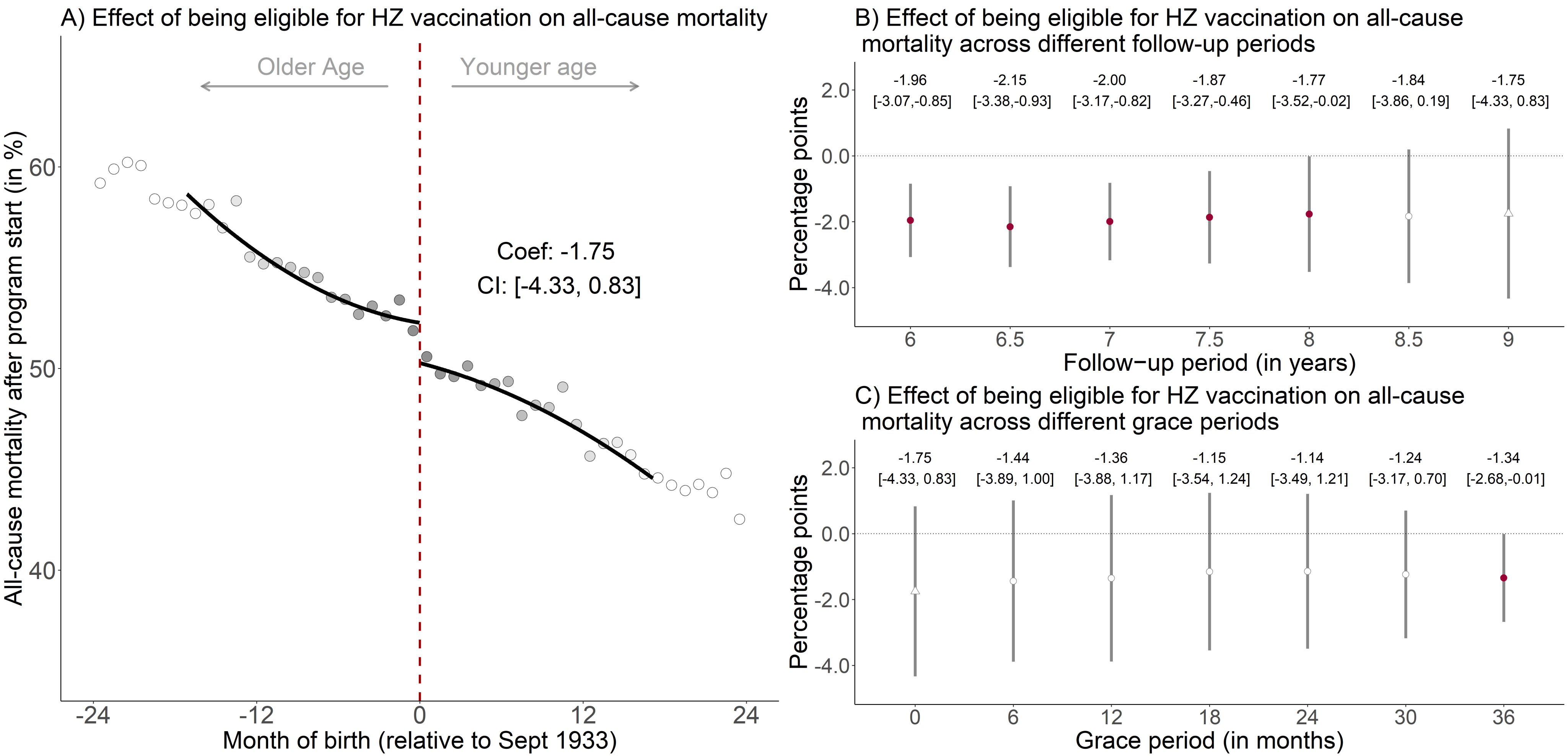


**Figure S16**. The effect of being eligible for herpes zoster vaccination on the percent of the population alive on September 1 2013 that died from any cause, using local squared instead of local linear regression.[1,2,3,4,5,6,7]

[1] Triangles (rather than points) depict our primary specification.

[2] Red (as opposed to white) fillings denote statistical significance ($p<0.05$).

[3] The grey shading of the dots is in proportion to the weight that observations from this birth-month received in the analysis.

[4] With “grace periods” we refer to time periods since September 1 2013 after which follow-up time is considered to begin to allow for the time needed for herpes zoster vaccination eligibility to begin affecting the outcome.

[5] Grey vertical bars depict 95% confidence intervals, which are estimated following Calonico, Cattaneo, and Titiunik (2014)(1).

[6] In Panel A, the sample size in the mean squared error-optimal bandwidth is 815,266 adults.
[7] The linear regression lines are drawn only in the mean squared error-optimal bandwidth.
Abbreviations: Coef=coefficient; CI=95% confidence interval; HZ=herpes zoster; Sept=September.

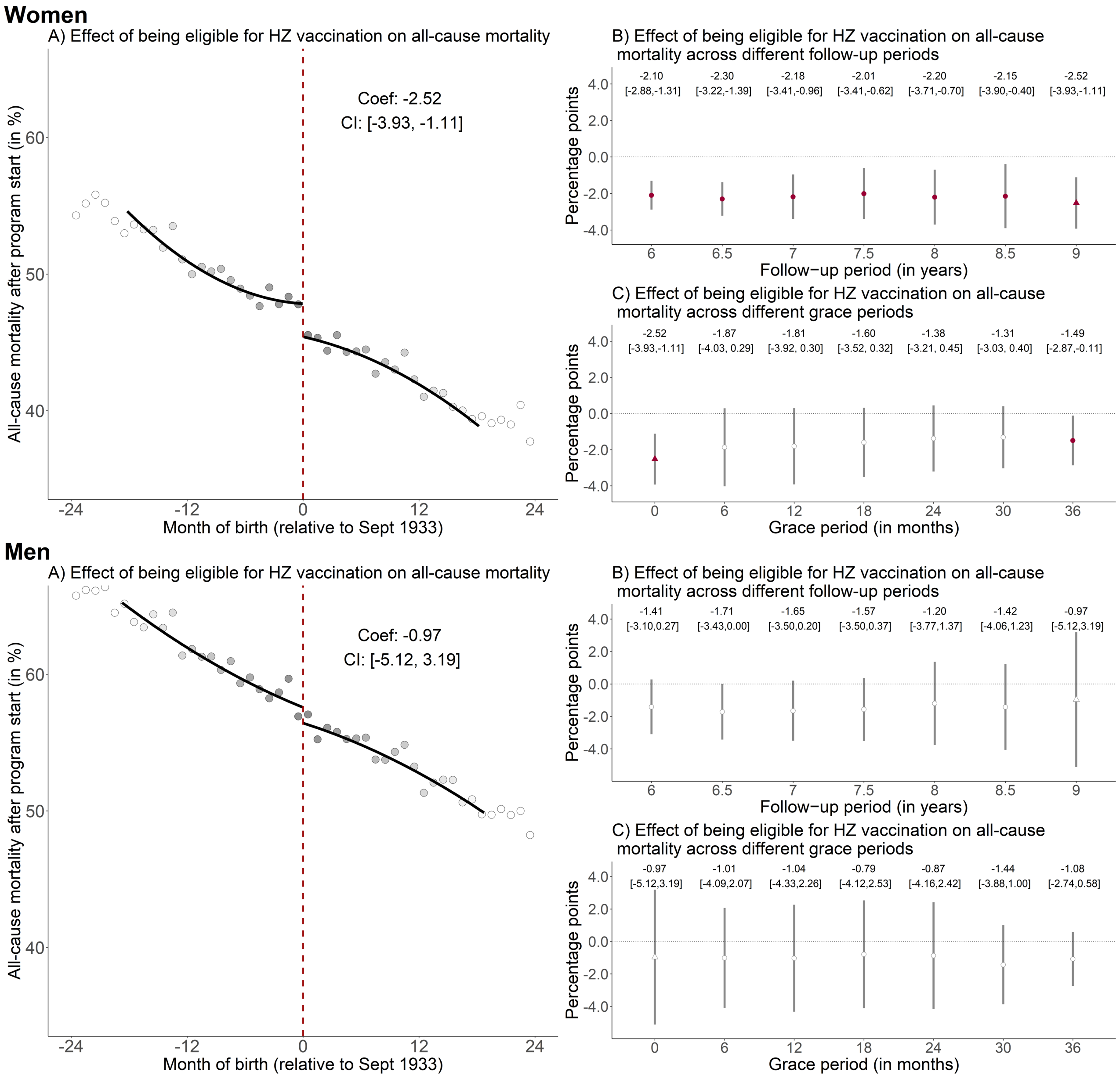

Women
A) Effect of being eligible for HZ vaccination on all-cause mortality
Coef: -2.52
CI: [-3.93, -1.11]
All-cause mortality after program start (in %)
Month of birth (relative to Sept 1933)
B) Effect of being eligible for HZ vaccination on all-cause mortality across different follow-up periods
-2.10 [-2.88,-1.31]
-2.30 [-3.22,-1.39]
-2.18 [-3.41,-0.96]
-2.01 [-3.41,-0.62]
-2.20 [-3.71,-0.70]
-2.15 [-3.90,-0.40]
-2.52 [-3.93,-1.11]
Percentage points
Follow−up period (in years)
C) Effect of being eligible for HZ vaccination on all-cause mortality across different grace periods
-2.52 [-3.93,-1.11]
-1.87 [-4.03, 0.29]
-1.81 [-3.92, 0.30]
-1.60 [-3.52, 0.32]
-1.38 [-3.21, 0.45]
-1.31 [-3.03, 0.40]
-1.49 [-2.87,-0.11]
Percentage points
Grace period (in months)
Men
A) Effect of being eligible for HZ vaccination on all-cause mortality
Coef: -0.97
CI: [-5.12, 3.19]
All-cause mortality after program start (in %)
Month of birth (relative to Sept 1933)
B) Effect of being eligible for HZ vaccination on all-cause mortality across different follow-up periods
-1.41 [-3.10,0.27]
-1.71 [-3.43,0.00]
-1.65 [-3.50,0.20]
-1.57 [-3.50,0.37]
-1.20 [-3.77,1.37]
-1.42 [-4.06,1.23]
-0.97 [-5.12,3.19]
Percentage points
Follow−up period (in years)
C) Effect of being eligible for HZ vaccination on all-cause mortality across different grace periods
-0.97 [-5.12,3.19]
-1.01 [-4.09,2.07]
-1.04 [-4.33,2.26]
-0.79 [-4.12,2.53]
-0.87 [-4.16,2.42]
-1.44 [-3.88,1.00]
-1.08 [-2.74,0.58]
Percentage points
Grace period (in months)

**Figure S17**. The effect of being eligible for herpes zoster vaccination on the percent of the population alive on September 1 2013 that died from any cause, using local squared instead of local linear regression and separately for women and men.[1,2,3,4,5,6,7]

[1] Triangles (rather than points) depict our primary specification.
[2] Red (as opposed to white) fillings denote statistical significance ($p<0.05$).
[3] The grey shading of the dots is in proportion to the weight that observations from this birth-month received in the analysis.
[4] With "grace periods" we refer to time periods since September 1 2013 after which follow-up time is considered to begin to allow for the time needed for herpes zoster vaccination eligibility to begin affecting the outcome.
[5] Grey vertical bars depict 95% confidence intervals, which are estimated following Calonico, Cattaneo, and Titiunik (2014)(1).
[6] In Panel A, the sample size in the mean squared error-optimal bandwidth is 550,680 and 440,831 for women and men, respectively.
[7] The linear regression lines are drawn only in the mean squared error-optimal bandwidth.
Abbreviations: Coef=coefficient; CI=95% confidence interval; HZ=herpes zoster; Sept=September.

| Shift of DOB threshold (months) | MSE-optimal BW | Coef [95% CI] (optimal BW) | Coef [95% CI] (BW=12 months) | Coef [95% CI] (BW=18 months) | Coef [95% CI] (BW=24 months) |
|---|---|---|---|---|---|
| -36 | 17.18 | -0.10 [-0.77, 0.56] | -0.13 [-0.95, 0.68] | -0.09 [-0.74, 0.56] | -0.05 [-0.69, 0.60] |
| -33 | 16.56 | 0.01 [-0.60, 0.62] | 0.07 [-0.62, 0.75] | 0.01 [-0.59, 0.61] | 0.05 [-0.57, 0.67] |
| -30 | 16.04 | 0.02 [-0.58, 0.63] | 0.06 [-0.65, 0.78] | 0.02 [-0.57, 0.61] | 0.04 [-0.57, 0.65] |
| -27 | 15.90 | 0.27 [-0.17, 0.72] | 0.31 [-0.13, 0.76] | 0.25 [-0.20, 0.71] | 0.20 [-0.33, 0.73] |
| -24 | 19.38 | -0.08 [-0.35, 0.19] | -0.22 [-0.47, 0.03] | -0.09 [-0.35, 0.17] | -0.06 [-0.36, 0.25] |
| -21 | 19.37 | -0.02 [-0.33, 0.30] | -0.09 [-0.39, 0.22] | -0.03 [-0.33, 0.28] | -0.00 [-0.35, 0.34] |
| -18 | 18.08 | -0.06 [-0.43, 0.31] | -0.03 [-0.43, 0.37] | -0.06 [-0.43, 0.31] | -0.02 [-0.43, 0.38] |
| -15 | 16.49 | -0.04 [-0.48, 0.41] | 0.00 [-0.52, 0.53] | -0.05 [-0.48, 0.39] | -0.04 [-0.50, 0.43] |
| -12 | 15.57 | 0.01 [-0.47, 0.50] | 0.00 [-0.56, 0.57] | 0.01 [-0.46, 0.47] | -0.04 [-0.53, 0.45] |
| -9 | 15.55 | 0.04 [-0.28, 0.36] | 0.02 [-0.28, 0.32] | 0.03 [-0.32, 0.38] | -0.05 [-0.47, 0.37] |
| -6 | 15.70 | -0.09 [-0.40, 0.22] | -0.11 [-0.43, 0.21] | -0.09 [-0.42, 0.24] | -0.13 [-0.54, 0.28] |
| -3 | 14.84 | -0.02 [-0.38, 0.33] | 0.03 [-0.34, 0.39] | -0.06 [-0.43, 0.31] | -0.07 [-0.52, 0.38] |
| 0 | 15.31 | -0.38 [-0.68, -0.08] | -0.38 [-0.66, -0.10] | -0.35 [-0.67, -0.02] | -0.26 [-0.66, 0.14] |
| 3 | 14.07 | 0.21 [-0.32, 0.74] | 0.24 [-0.33, 0.80] | 0.18 [-0.34, 0.69] | 0.14 [-0.42, 0.70] |
| 6 | 14.81 | 0.18 [-0.34, 0.71] | 0.13 [-0.45, 0.70] | 0.20 [-0.31, 0.71] | 0.17 [-0.38, 0.72] |
| 9 | 15.18 | 0.28 [-0.20, 0.77] | 0.27 [-0.28, 0.82] | 0.28 [-0.19, 0.75] | 0.26 [-0.25, 0.77] |
| 12 | 17.03 | -0.07 [-0.36, 0.21] | -0.13 [-0.42, 0.15] | -0.06 [-0.35, 0.23] | 0.02 [-0.33, 0.37] |
| 15 | 18.06 | -0.26 [-0.60, 0.07] | -0.32 [-0.70, 0.06] | -0.27 [-0.60, 0.07] | -0.16 [-0.52, 0.19] |
| 18 | 17.25 | -0.00 [-0.38, 0.38] | 0.05 [-0.42, 0.53] | -0.01 [-0.39, 0.36] | -0.02 [-0.41, 0.36] |
| 21 | 17.64 | 0.15 [-0.14, 0.44] | 0.23 [-0.06, 0.52] | 0.14 [-0.15, 0.43] | 0.07 [-0.27, 0.40] |
| 24 | 18.55 | -0.02 [-0.26, 0.23] | -0.08 [-0.29, 0.14] | -0.02 [-0.26, 0.22] | -0.04 [-0.33, 0.25] |
| 27 | 18.60 | -0.15 [-0.45, 0.16] | -0.25 [-0.60, 0.10] | -0.16 [-0.46, 0.15] | -0.13 [-0.45, 0.20] |
| 30 | 18.28 | -0.05 [-0.35, 0.26] | 0.01 [-0.33, 0.34] | -0.05 [-0.35, 0.26] | -0.02 [-0.35, 0.31] |
| 33 | 17.78 | 0.01 [-0.23, 0.24] | 0.03 [-0.21, 0.27] | 0.00 [-0.23, 0.24] | 0.01 [-0.27, 0.29] |
| 36 | 17.99 | 0.03 [-0.15, 0.21] | -0.01 [-0.17, 0.14] | 0.03 [-0.15, 0.21] | 0.01 [-0.23, 0.25] |

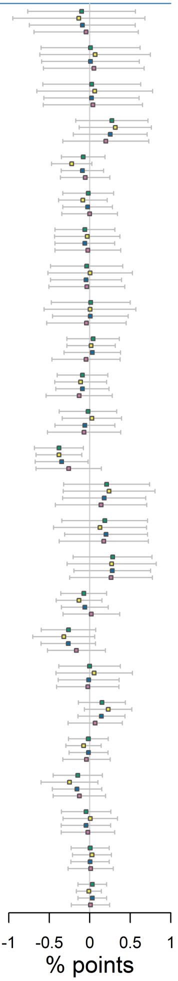


**Figure S18**. The effect of being eligible for herpes zoster vaccination on the percent of the population alive on September 1 2013 that died due to dementia, across different bandwidth choices and shifts of the date-of-birth threshold.[1,2]

[1] The bandwidth is the window (in months) in participants' date of birth that is drawn around the September 2 1933 eligibility threshold.

[2] When shifting the date-of-birth threshold, we only shifted the date-of-birth threshold in our analysis but not the start and end date of the follow-up period. For example, when moving the start date of the program to “-18”, we started the follow-up period on September 1 2013 and compared individuals around the March 2 1932 date-of-birth threshold.

Abbreviations: DOB=date of birth; MSE=mean squared error; Coef.=coefficient; CI=95% confidence interval; BW=bandwidth.

| Shift of DOB threshold (months) | MSE-optimal BW | Coef [95% CI] (optimal BW) | Coef [95% CI] (BW=12 months) | Coef [95% CI] (BW=18 months) | Coef [95% CI] (BW=24 months) |
|---|---|---|---|---|---|
| -36 | 16.05 | -0.58 [-2.74, 1.58] | -0.74 [-3.30, 1.82] | -0.52 [-2.60, 1.56] | -0.48 [-2.61, 1.66] |
| -33 | 14.17 | 0.29 [-1.37, 1.96] | 0.42 [-1.25, 2.09] | 0.21 [-1.60, 2.01] | 0.13 [-2.10, 2.36] |
| -30 | 14.70 | 0.07 [-1.36, 1.50] | 0.06 [-1.40, 1.52] | -0.00 [-1.56, 1.55] | 0.03 [-1.94, 2.00] |
| -27 | 13.88 | 0.93 [-0.93, 2.79] | 1.02 [-0.89, 2.94] | 0.79 [-1.08, 2.66] | 0.62 [-1.54, 2.79] |
| -24 | 18.11 | -0.91 [-2.14, 0.33] | -1.56 [-2.91, -0.20] | -0.92 [-2.15, 0.32] | -0.68 [-1.98, 0.62] |
| -21 | 15.92 | -0.05 [-1.65, 1.56] | -0.06 [-1.73, 1.62] | -0.03 [-1.65, 1.60] | 0.01 [-1.80, 1.82] |
| -18 | 15.47 | 0.34 [-1.11, 1.79] | 0.49 [-1.08, 2.06] | 0.23 [-1.26, 1.72] | 0.23 [-1.52, 1.98] |
| -15 | 16.53 | -0.00 [-1.40, 1.39] | 0.08 [-1.44, 1.59] | 0.00 [-1.38, 1.39] | 0.02 [-1.52, 1.56] |
| -12 | 16.88 | -0.46 [-2.17, 1.26] | -0.74 [-2.92, 1.44] | -0.43 [-2.10, 1.23] | -0.39 [-2.01, 1.23] |
| -9 | 16.08 | 0.20 [-0.87, 1.27] | 0.25 [-0.73, 1.23] | 0.22 [-0.93, 1.37] | 0.08 [-1.33, 1.49] |
| -6 | 16.23 | 0.27 [-0.85, 1.39] | 0.42 [-0.84, 1.67] | 0.20 [-0.95, 1.34] | 0.10 [-1.28, 1.48] |
| -3 | 16.41 | 0.27 [-0.86, 1.41] | 0.41 [-0.78, 1.59] | 0.21 [-0.94, 1.36] | 0.13 [-1.19, 1.46] |
| 0 | 17.12 | -1.23 [-2.35, -0.11] | -1.62 [-2.93, -0.32] | -1.16 [-2.27, -0.05] | -0.93 [-2.11, 0.24] |
| 3 | 15.38 | 0.45 [-0.84, 1.74] | 0.63 [-0.78, 2.04] | 0.37 [-0.93, 1.67] | 0.22 [-1.26, 1.71] |
| 6 | 15.71 | 0.59 [-0.62, 1.80] | 0.64 [-0.63, 1.92] | 0.48 [-0.76, 1.71] | 0.37 [-1.07, 1.81] |
| 9 | 16.11 | 0.65 [-0.84, 2.14] | 0.71 [-0.94, 2.36] | 0.60 [-0.86, 2.07] | 0.51 [-1.04, 2.05] |
| 12 | 17.74 | -0.81 [-1.99, 0.36] | -1.32 [-2.74, 0.09] | -0.79 [-1.97, 0.38] | -0.52 [-1.72, 0.69] |
| 15 | 16.88 | -0.34 [-1.44, 0.75] | -0.35 [-1.48, 0.78] | -0.30 [-1.41, 0.82] | -0.11 [-1.36, 1.13] |
| 18 | 15.42 | 0.42 [-0.65, 1.48] | 0.48 [-0.65, 1.62] | 0.31 [-0.80, 1.41] | 0.29 [-1.03, 1.61] |
| 21 | 13.46 | 0.73 [-0.48, 1.95] | 0.71 [-0.52, 1.93] | 0.61 [-0.69, 1.91] | 0.41 [-1.19, 2.01] |
| 24 | 15.35 | -0.78 [-2.01, 0.45] | -1.02 [-2.41, 0.37] | -0.63 [-1.83, 0.57] | -0.59 [-1.91, 0.73] |
| 27 | 15.30 | -0.25 [-1.36, 0.86] | -0.19 [-1.34, 0.97] | -0.28 [-1.43, 0.86] | -0.33 [-1.68, 1.02] |
| 30 | 14.80 | 0.30 [-0.68, 1.29] | 0.56 [-0.48, 1.61] | 0.10 [-0.94, 1.13] | -0.05 [-1.32, 1.23] |
| 33 | 16.34 | -0.13 [-0.93, 0.67] | -0.12 [-0.93, 0.68] | -0.18 [-0.99, 0.64] | -0.22 [-1.20, 0.77] |
| 36 | 16.31 | -0.42 [-1.49, 0.65] | -0.64 [-1.87, 0.60] | -0.34 [-1.40, 0.71] | -0.28 [-1.42, 0.85] |

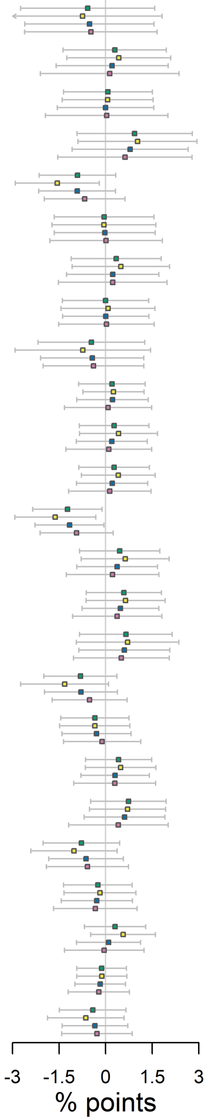


■ optimal BW ■ BW=12 months ■ BW=18 months ■ BW=24 months

**Figure S19**. The effect of being eligible for herpes zoster vaccination on the percent of the population alive on September 1 2013 that died from any cause, across different bandwidth choices and shifts of the date-of-birth threshold.[1]

[1] The bandwidth is the window (in months) in participants' date of birth that is drawn around the September 2 1933 eligibility threshold.

[2] When shifting the date-of-birth threshold, we only shifted the date-of-birth threshold in our analysis but not the start and end date of the follow-up period. For example, when moving the start date of the program to "-18", we started the follow-up period on September 1 2013 and compared individuals around the March 2 1932 date-of-birth threshold.

Abbreviations: DOB=date of birth; MSE=mean squared error; Coef.=coefficient; CI=95% confidence interval; BW=bandwidth.

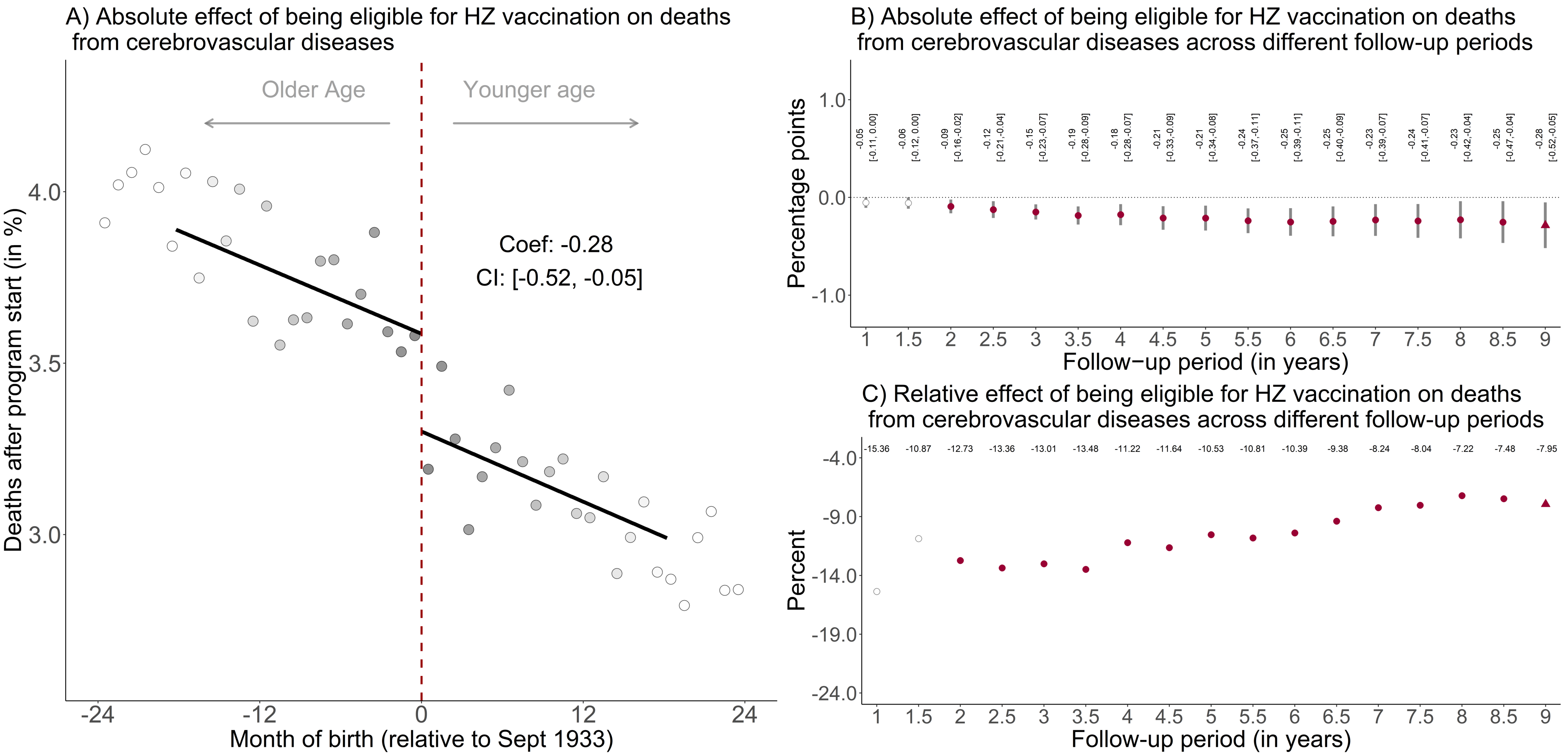


**Figure S20.** The effect of being eligible for herpes zoster vaccination on the percent of the population alive on September 1 2013 that died due to cerebrovascular diseases.[1,2,3,4,5,6]

[1] Triangles (rather than points) depict our primary specification.

[2] Red (as opposed to white) fillings denote statistical significance ($p<0.05$).

[3] The grey shading of the dots is in proportion to the weight that observations from this birth-month received in the analysis.

[4] Grey vertical bars depict 95% confidence intervals.
[5] In Panel A, the sample size in the mean squared error-optimal bandwidth is 991,511 adults.
[6] The linear regression lines are drawn only in the mean squared error-optimal bandwidth.
Abbreviations: Coef=coefficient; CI=95% confidence interval; HZ=herpes zoster; Sept=September.

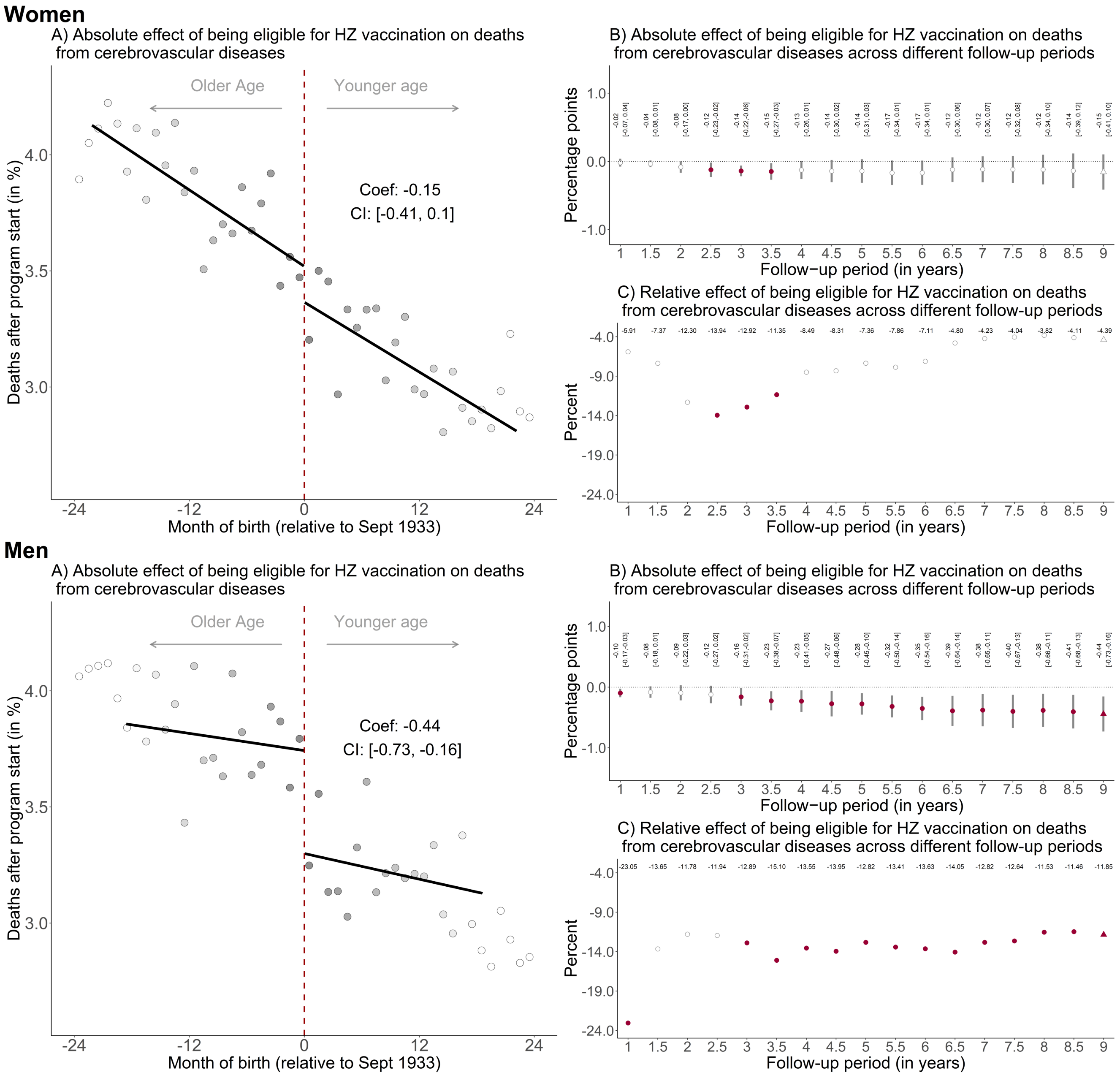
Women
A) Absolute effect of being eligible for HZ vaccination on deaths from cerebrovascular diseases
Older Age
Younger age
Coef: -0.15
CI: [-0.41, 0.1]
Deaths after program start (in %)
Month of birth (relative to Sept 1933)
B) Absolute effect of being eligible for HZ vaccination on deaths from cerebrovascular diseases across different follow-up periods
Percentage points
Follow−up period (in years)
C) Relative effect of being eligible for HZ vaccination on deaths from cerebrovascular diseases across different follow-up periods
Percent
Follow-up period (in years)
Men
A) Absolute effect of being eligible for HZ vaccination on deaths from cerebrovascular diseases
Older Age
Younger age
Coef: -0.44
CI: [-0.73, -0.16]
Deaths after program start (in %)
Month of birth (relative to Sept 1933)
B) Absolute effect of being eligible for HZ vaccination on deaths from cerebrovascular diseases across different follow-up periods
Percentage points
Follow−up period (in years)
C) Relative effect of being eligible for HZ vaccination on deaths from cerebrovascular diseases across different follow-up periods
Percent
Follow-up period (in years)

**Figure S21.** The effect of being eligible for herpes zoster vaccination on the percent of the population alive on September 1 2013 that died due to cerebrovascular diseases, separately for women and men.[1,2,3,4,5,6]

[1] Triangles (rather than points) depict our primary specification.
[2] Red (as opposed to white) fillings denote statistical significance ($p<0.05$).
[3] The grey shading of the dots is in proportion to the weight that observations from this birth-month received in the analysis.
[4] Grey vertical bars depict 95% confidence intervals.
[5] In Panel A, the sample size in the mean squared error-optimal bandwidth is 643,491 and 440,831 for women and men, respectively.
[6] The linear regression lines are drawn only in the mean squared error-optimal bandwidth.
Abbreviations: Coef=coefficient; CI=95% confidence interval; HZ=herpes zoster; Sept=September.

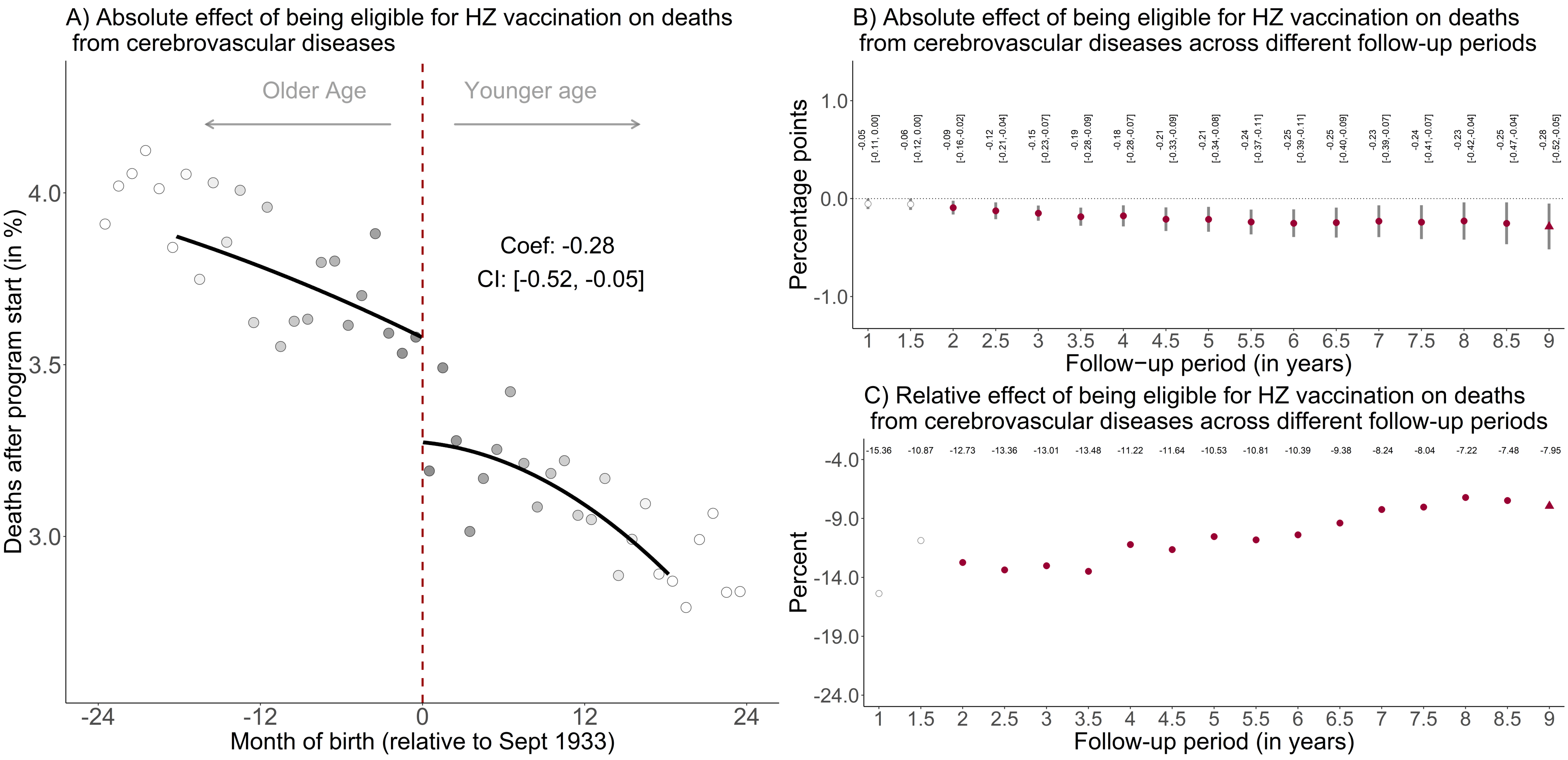


**Figure S22**. The effect of being eligible for herpes zoster vaccination on the percent of the population alive on September 1 2013 that died from cerebrovascular diseases, using local squared instead of local linear regression.[1,2,3,4,5,6]

[1] Triangles (rather than points) depict our primary specification.

[2] Red (as opposed to white) fillings denote statistical significance (p<0.05).

[3] The grey shading of the dots is in proportion to the weight that observations from this birth-month received in the analysis.

[4] Grey vertical bars depict 95% confidence intervals, which are estimated following Calonico, Cattaneo, and Titiunik (2014)(1).

[5] In Panel A, the sample size in the mean squared error-optimal bandwidth is 991,511 adults.

[6] The linear regression lines are drawn only in the mean squared error-optimal bandwidth.
Abbreviations: Coef=coefficient; CI=95% confidence interval; HZ=herpes zoster; Sept=September.

### Women

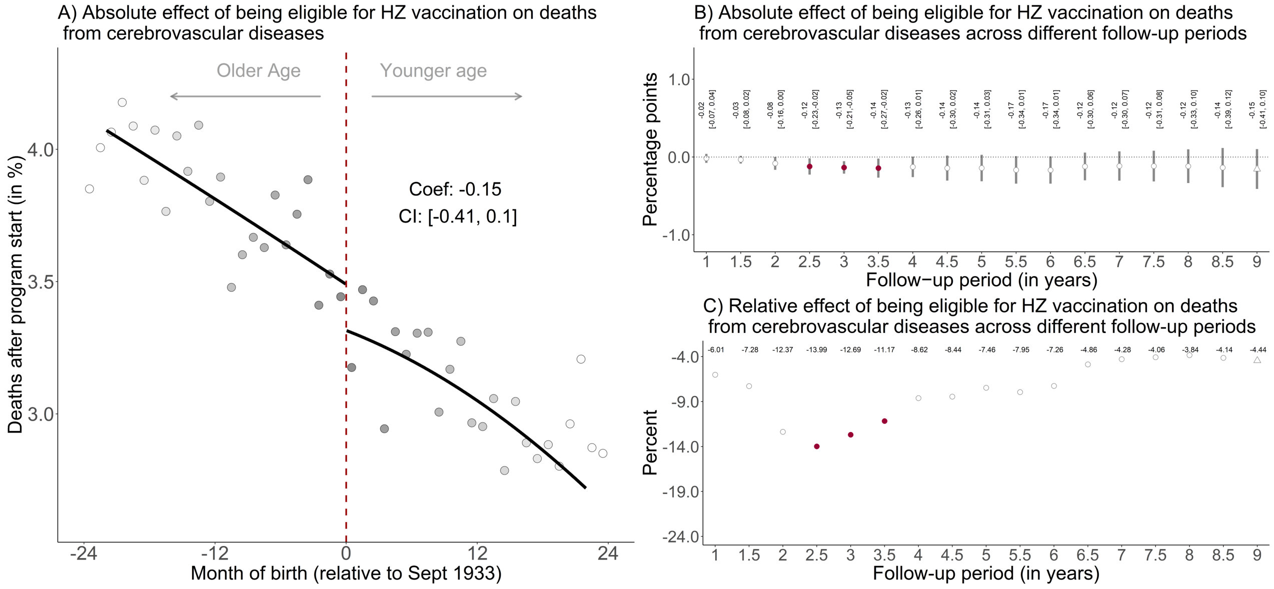

A) Absolute effect of being eligible for HZ vaccination on deaths from cerebrovascular diseases
Older Age
Younger age
Coef: -0.15
CI: [-0.41, 0.1]
Deaths after program start (in %)
Month of birth (relative to Sept 1933)
B) Absolute effect of being eligible for HZ vaccination on deaths from cerebrovascular diseases across different follow-up periods
Percentage points
Follow−up period (in years)
C) Relative effect of being eligible for HZ vaccination on deaths from cerebrovascular diseases across different follow-up periods
-6.01 -7.28 -12.37 -13.99 -12.69 -11.17 -8.62 -8.44 -7.46 -7.95 -7.26 -4.86 -4.28 -4.06 -3.84 -4.14 -4.44
Percent
Follow-up period (in years)


### Men

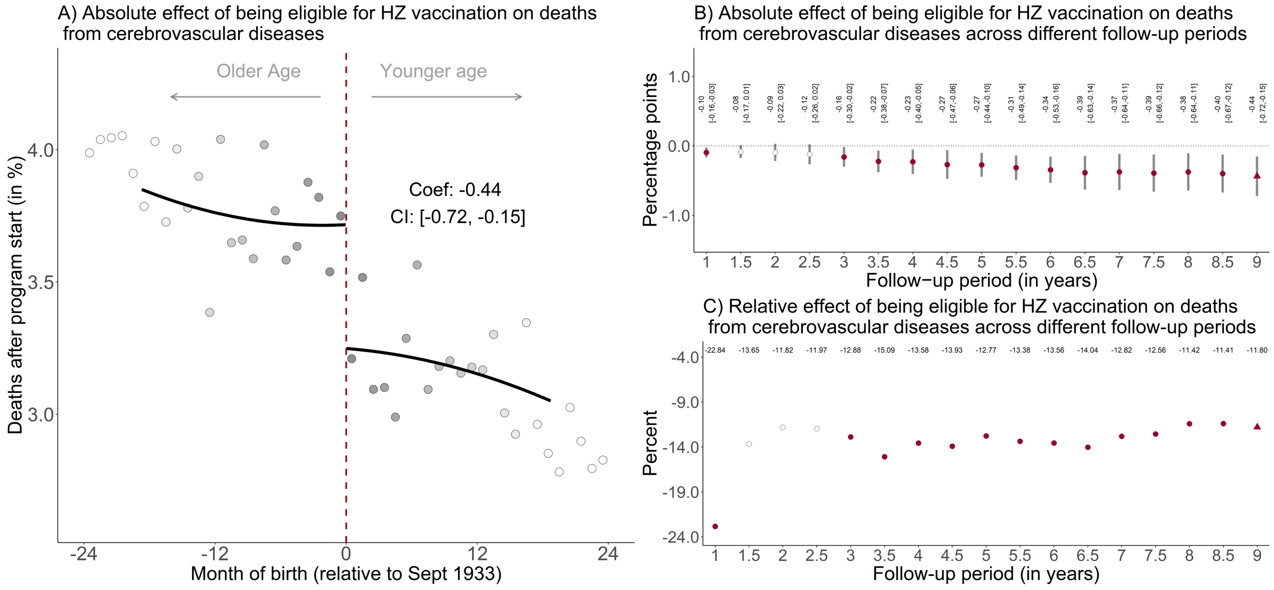

A) Absolute effect of being eligible for HZ vaccination on deaths from cerebrovascular diseases
Older Age
Younger age
Coef: -0.44
CI: [-0.72, -0.15]
Deaths after program start (in %)
Month of birth (relative to Sept 1933)
B) Absolute effect of being eligible for HZ vaccination on deaths from cerebrovascular diseases across different follow-up periods
Percentage points
Follow−up period (in years)
C) Relative effect of being eligible for HZ vaccination on deaths from cerebrovascular diseases across different follow-up periods
-22.84 -13.65 -11.82 -11.97 -12.88 -15.09 -13.58 -13.93 -12.77 -13.38 -13.56 -14.04 -12.82 -12.56 -11.42 -11.41 -11.80
Percent
Follow-up period (in years)

**Figure S23**. The effect of being eligible for herpes zoster vaccination on the percent of the population alive on September 1 2013 that died from cerebrovascular diseases, using local squared instead of local linear regression and separately for women and men.[1,2,3,4,5,6]

[1] Triangles (rather than points) depict our primary specification.
[2] Red (as opposed to white) fillings denote statistical significance ($p<0.05$).
[3] The grey shading of the dots is in proportion to the weight that observations from this birth-month received in the analysis.
[4] Grey vertical bars depict 95% confidence intervals, which are estimated following Calonico, Cattaneo, and Titiunik (2014)(1).
[5] In Panel A, the sample size in the mean squared error-optimal bandwidth is 643,491 and 440,831 among women and men, respectively.
[6] The linear regression lines are drawn only in the mean squared error-optimal bandwidth.
Abbreviations: Coef=coefficient; CI=95% confidence interval; HZ=herpes zoster; Sept=September.

| Shift of DOB threshold (months) | MSE-optimal BW | Coef [95% CI] (optimal BW) | Coef [95% CI] (BW=12 months) | Coef [95% CI] (BW=18 months) | Coef [95% CI] (BW=24 months) |
|---|---|---|---|---|---|
| -36 | 23.89 | -0.12 [-0.39, 0.16] | -0.08 [-0.48, 0.31] | -0.10 [-0.41, 0.21] | -0.12 [-0.39, 0.15] |
| -33 | 22.38 | -0.19 [-0.44, 0.05] | -0.19 [-0.46, 0.09] | -0.18 [-0.42, 0.06] | -0.19 [-0.43, 0.05] |
| -30 | 15.45 | 0.06 [-0.23, 0.36] | 0.10 [-0.20, 0.40] | 0.03 [-0.28, 0.33] | -0.03 [-0.39, 0.34] |
| -27 | 14.44 | 0.10 [-0.20, 0.40] | 0.12 [-0.19, 0.43] | 0.08 [-0.23, 0.39] | 0.02 [-0.35, 0.40] |
| -24 | 18.17 | -0.11 [-0.31, 0.09] | -0.19 [-0.39, 0.01] | -0.11 [-0.31, 0.08] | -0.11 [-0.34, 0.12] |
| -21 | 15.75 | 0.07 [-0.15, 0.29] | 0.09 [-0.11, 0.28] | 0.06 [-0.18, 0.29] | 0.06 [-0.24, 0.35] |
| -18 | 16.72 | 0.06 [-0.19, 0.30] | 0.10 [-0.18, 0.37] | 0.06 [-0.19, 0.30] | 0.09 [-0.19, 0.37] |
| -15 | 22.85 | 0.05 [-0.16, 0.26] | -0.03 [-0.29, 0.23] | 0.00 [-0.22, 0.22] | 0.05 [-0.16, 0.26] |
| -12 | 28.43 | 0.08 [-0.14, 0.31] | -0.01 [-0.41, 0.40] | 0.05 [-0.25, 0.35] | 0.08 [-0.17, 0.32] |
| -9 | 31.78 | 0.13 [-0.04, 0.30] | 0.17 [-0.06, 0.39] | 0.17 [-0.03, 0.36] | 0.14 [-0.03, 0.32] |
| -6 | 24.75 | 0.04 [-0.13, 0.22] | 0.07 [-0.12, 0.26] | 0.06 [-0.11, 0.23] | 0.04 [-0.13, 0.22] |
| -3 | 17.58 | -0.19 [-0.41, 0.04] | -0.22 [-0.46, 0.02] | -0.18 [-0.41, 0.04] | -0.16 [-0.41, 0.08] |
| 0 | 18.22 | -0.28 [-0.52, -0.05] | -0.30 [-0.55, -0.06] | -0.29 [-0.52, -0.05] | -0.26 [-0.51, -0.01] |
| 3 | 18.87 | -0.12 [-0.36, 0.12] | -0.07 [-0.35, 0.21] | -0.12 [-0.36, 0.12] | -0.13 [-0.38, 0.11] |
| 6 | 33.45 | 0.07 [-0.10, 0.23] | 0.24 [-0.00, 0.49] | 0.17 [-0.03, 0.38] | 0.11 [-0.07, 0.29] |
| 9 | 30.67 | 0.07 [-0.10, 0.23] | 0.05 [-0.19, 0.28] | 0.09 [-0.09, 0.28] | 0.09 [-0.08, 0.25] |
| 12 | 29.70 | 0.06 [-0.09, 0.20] | -0.05 [-0.24, 0.13] | 0.01 [-0.15, 0.17] | 0.06 [-0.09, 0.21] |
| 15 | 27.58 | 0.09 [-0.07, 0.24] | 0.04 [-0.19, 0.28] | 0.04 [-0.14, 0.22] | 0.08 [-0.08, 0.24] |
| 18 | 21.38 | 0.03 [-0.15, 0.20] | 0.02 [-0.17, 0.22] | 0.03 [-0.14, 0.21] | 0.04 [-0.13, 0.22] |
| 21 | 15.14 | 0.10 [-0.11, 0.32] | 0.13 [-0.10, 0.36] | 0.09 [-0.13, 0.30] | 0.05 [-0.20, 0.30] |
| 24 | 21.68 | -0.12 [-0.26, 0.02] | -0.15 [-0.31, 0.00] | -0.13 [-0.27, 0.01] | -0.12 [-0.26, 0.03] |
| 27 | 25.15 | -0.10 [-0.22, 0.03] | -0.08 [-0.23, 0.07] | -0.09 [-0.22, 0.03] | -0.09 [-0.22, 0.03] |
| 30 | 28.33 | -0.05 [-0.16, 0.06] | 0.02 [-0.12, 0.17] | -0.02 [-0.13, 0.10] | -0.04 [-0.15, 0.07] |
| 33 | 14.22 | 0.14 [-0.06, 0.34] | 0.13 [-0.07, 0.34] | 0.10 [-0.10, 0.31] | 0.06 [-0.18, 0.30] |
| 36 | 14.77 | 0.02 [-0.22, 0.26] | 0.01 [-0.26, 0.29] | 0.03 [-0.20, 0.27] | 0.02 [-0.23, 0.28] |

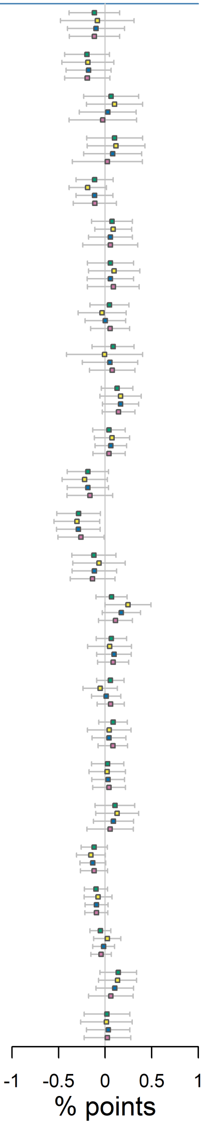


optimal BW   BW=12 months   BW=18 months   BW=24 months

**Figure S24**. The effect of being eligible for herpes zoster vaccination on the percent of the population alive on September 1 2013 that died from cerebrovascular diseases, across different bandwidth choices and shifts of the date-of-birth threshold.[1]

[1] The bandwidth is the window (in months) in participants' date of birth that is drawn around the September 2 1933 eligibility threshold.

[2] When shifting the date-of-birth threshold, we only shifted the date-of-birth threshold in our analysis but not the start and end date of the follow-up period. For example, when moving the start date of the program to "-18", we started the follow-up period on September 1 2013 and compared individuals around the March 2 1932 date-of-birth threshold.

Abbreviations: DOB=date of birth; MSE=mean squared error; Coef.=coefficient; CI=95% confidence interval; BW=bandwidth.

| Variable | Total | | Women | | Men | |
|---|---|---|---|---|---|---|
| | N | % | N | % | N | % |
| Population in Sept 2013 | 5,077,426 | 100 | 2,824,529 | 55.6 | 2,252,897 | 44.4 |
| Population in Sept 2004 | 6,798,800 | 100 | 3,616,753 | 53.2 | 3,182,047 | 46.8 |
| *Deaths from Sept 1 2004 to Aug 31 2013* | | | | | | |
| All-cause mortality | 1,721,374 | 25.5 | 792,224 | 22.1 | 929,150 | 29.5 |
| Ischemic heart diseases (I20–I25) | 276,903 | 4.1 | 102,096 | 2.8 | 174,807 | 5.5 |
| Malignant neoplasm of trachea, bronchus, and lung (C33-C34) | 140,304 | 2.1 | 58,791 | 1.6 | 81,513 | 2.6 |
| Cerebrovascular diseases (I60–I69) | 133,599 | 2 | 69,659 | 1.9 | 63,940 | 2 |
| Chronic lower respiratory diseases (J40–J47) | 120,233 | 1.8 | 57,567 | 1.6 | 62,666 | 2 |
| Dementia of any type (F01, F03, G30) | 75,770 | 1.1 | 44,893 | 1.3 | 30,877 | 1 |
| Influenza and pneumonia (J09–J18) | 66,615 | 1 | 32,888 | 0.9 | 33,727 | 1.1 |
| Malignant neoplasm of colon, sigmoid, rectum, and anus (C18–C21) | 59,739 | 0.9 | 25,191 | 0.7 | 34,548 | 1.1 |
| Malignant neoplasms, stated or presumed to be primary of lymphoid, hematopoietic, and related tissue (C81–C96) | 47,292 | 0.7 | 20,346 | 0.6 | 26,946 | 0.9 |
| Dementia of unspecified type (F03) | 44,155 | 0.7 | 26,499 | 0.7 | 17,656 | 0.6 |
| Malignant neoplasm of prostate (C61) | 42,821 | 0.6 | 0 | 0 | 42,821 | 1.4 |
| Malignant neoplasm of breast (C50) | 33,580 | 0.5 | 33,300 | 0.9 | 280 | 0 |
| Aortic aneurysm and dissection (I71) | 32,459 | 0.5 | 11,671 | 0.3 | 20,788 | 0.7 |
| Malignant neoplasm of pancreas (C25) | 30,903 | 0.5 | 15,489 | 0.4 | 15,414 | 0.5 |
| Diseases of the urinary system (N00–N39) | 30,154 | 0.4 | 15,952 | 0.4 | 14,202 | 0.5 |
| Malignant neoplasm of esophagus (C15) | 27,648 | 0.4 | 8,984 | 0.3 | 18,664 | 0.6 |
| Heart failure and complications, and ill-defined heart disease (I50–I51) | 24,568 | 0.4 | 12,173 | 0.3 | 12,395 | 0.4 |
| Alzheimer’s disease (G30) | 21,225 | 0.3 | 12,932 | 0.4 | 8,293 | 0.3 |
| Malignant neoplasm of stomach (C16) | 20,295 | 0.3 | 6,785 | 0.2 | 13,510 | 0.4 |
| Parkinson’s disease (G20) | 19,932 | 0.3 | 7,158 | 0.2 | 12,774 | 0.4 |
| Diabetes (E10–E14) | 19,584 | 0.3 | 9,320 | 0.3 | 10,264 | 0.3 |
| Malignant neoplasm of bladder (C67) | 19,152 | 0.3 | 5,646 | 0.2 | 13,506 | 0.4 |
| Pulmonary edema and other interstitial pulmonary diseases (I26–I28) | 18,834 | 0.3 | 6,680 | 0.2 | 12,154 | 0.4 |
| Malignant neoplasm of ovary (C56) | 14,970 | 0.2 | 14,970 | 0.4 | 0 | 0 |
| Cirrhosis and other diseases of liver (K70–K76) | 14,813 | 0.2 | 6,359 | 0.2 | 8,454 | 0.3 |
| Nonrheumatic valve disorders and endocarditis (I34–I38) | 14,657 | 0.2 | 7,143 | 0.2 | 7,514 | 0.2 |
| Malignant neoplasm of liver and intrahepatic bile ducts (C22) | 14,109 | 0.2 | 5,587 | 0.2 | 8,522 | 0.3 |
| Hypertensive diseases (I10–I15) | 13,746 | 0.2 | 6,980 | 0.2 | 6,766 | 0.2 |
| Diseases of the musculoskeletal system and connective tissue (M00–M99) | 13,673 | 0.2 | 8,649 | 0.2 | 5,024 | 0.2 |
| Malignant neoplasm of kidney, except renal pelvis (C64) | 13,319 | 0.2 | 5,089 | 0.1 | 8,230 | 0.3 |
| In situ and benign neoplasms, and neoplasms of uncertain or unknown behavior (D00–D48) | 12,131 | 0.2 | 5,598 | 0.2 | 6,533 | 0.2 |
| Pulmonary heart disease and diseases of pulmonary circulation (I26–I28) | 11,767 | 0.2 | 6,825 | 0.2 | 4,942 | 0.2 |
| Malignant neoplasm of brain (C71) | 10,876 | 0.2 | 4,710 | 0.1 | 6,166 | 0.2 |
| Accidental falls (W00–W19) | 10,549 | 0.2 | 4,813 | 0.1 | 5,736 | 0.2 |
| Vascular Dementia (F01) | 10,390 | 0.2 | 5,462 | 0.2 | 4,928 | 0.2 |
| Cardiac arrhythmias (I47–I49) | 10,304 | 0.2 | 5,998 | 0.2 | 4,306 | 0.1 |
| Appendicitis, hernia and intestinal obstruction (K35–K46, K56) | 10,011 | 0.1 | 5,136 | 0.1 | 4,875 | 0.2 |
| Systemic atrophies primarily affecting the central nervous system (G10–G12) | 9,513 | 0.1 | 4,397 | 0.1 | 5,116 | 0.2 |
| Malignant neoplasm of uterus (C53–C55) | 9,042 | 0.1 | 9,042 | 0.3 | 0 | 0 |

| | | | | | | |
|---|---|---|---|---|---|---|
| Melanoma and other malignant neoplasms of skin (C43-C44) | 7,967 | 0.1 | 3,123 | 0.1 | 4,844 | 0.2 |
| Septicemia (A40–A41) | 7,407 | 0.1 | 3,932 | 0.1 | 3,475 | 0.1 |
| Intestinal infectious diseases (A00–A09) | 6,714 | 0.1 | 3,947 | 0.1 | 2,767 | 0.1 |
| Symptoms, signs, and ill-defined conditions (R00-R99) | 6,499 | 0.1 | 3,937 | 0.1 | 2,562 | 0.1 |
| Acute respiratory infections other than influenza and pneumonia (J00–J06, J20–J22) | 4,625 | 0.1 | 2,379 | 0.1 | 2,246 | 0.1 |
| Chronic rheumatic heart diseases (I05–I09) | 4,485 | 0.1 | 3,006 | 0.1 | 1,479 | 0 |
| Suicide and injury/poisoning of undetermined intent (X60–X84, Y10–Y34) | 4,305 | 0.1 | 1,312 | 0 | 2,993 | 0.1 |
| Cardiomyopathy (I42) | 3,883 | 0.1 | 1,294 | 0 | 2,589 | 0.1 |
| Land transport accidents (V01–V89) | 2,677 | 0 | 1,061 | 0 | 1,616 | 0.1 |
| Malignant neoplasm of gallbladder and other parts of biliary tract (C23-C24) | 2,655 | 0 | 1,717 | 0 | 938 | 0 |
| Malignant neoplasm of larynx (C32) | 2,633 | 0 | 506 | 0 | 2,127 | 0.1 |
| Epilepsy and status epilepticus (G40-G41) | 1,825 | 0 | 922 | 0 | 903 | 0 |
| Congenital malformations, deformations, and chromosomal abnormalities (Q00–Q99) | 1,529 | 0 | 800 | 0 | 729 | 0 |
| Tuberculosis (A15–A19, B90) | 1,317 | 0 | 563 | 0 | 754 | 0 |
| Atherosclerosis (I70) | 1,108 | 0 | 526 | 0 | 582 | 0 |
| Accidental threats to breathing (W75–W84) | 1,087 | 0 | 490 | 0 | 597 | 0 |
| Cerebral palsy and other paralytic syndromes (G80–G83) | 827 | 0 | 321 | 0 | 506 | 0 |
| Disorders of fluid, electrolyte, and acid (E86–E87) | 752 | 0 | 433 | 0 | 319 | 0 |
| Mental and behavioral disorders due to psychoactive substance use (F10–F19) | 701 | 0 | 198 | 0 | 503 | 0 |
| Malignant neoplasms of bone and articular cartilage (C40–C41) | 625 | 0 | 254 | 0 | 371 | 0 |
| Accidental poisoning (X40–X49) | 545 | 0 | 288 | 0 | 257 | 0 |
| Meningitis and meningococcal infection (A39) | 465 | 0 | 254 | 0 | 211 | 0 |
| Vaccine-preventable diseases (A33–A37, A49.2, A80, B01, B02, B05, B06, B15, B16, B17.0, B18.0, B18.1, B26, B91, G14) | 409 | 0 | 191 | 0 | 218 | 0 |
| Homicide and probable homicide (U50.9, X85–Y09, Y87.1) | 356 | 0 | 173 | 0 | 183 | 0 |
| Malnutrition, nutritional anemias, and other nutritional deficiencies (D50–D53, E40–E64) | 345 | 0 | 206 | 0 | 139 | 0 |
| Accidental drowning and submersion (W65–W74) | 226 | 0 | 76 | 0 | 150 | 0 |
| HIV (B20–B24) | 129 | 0 | 19 | 0 | 110 | 0 |
| Cardiac arrest (I46) | 87 | 0 | 52 | 0 | 35 | 0 |
| Respiratory failure (J96) | 66 | 0 | 40 | 0 | 26 | 0 |
| Vector-borne diseases and rabies (A20, A44, A75–A79, A82–A84, A85.2, A90–A98, B50–B57) | 11 | 0 | 4 | 0 | 7 | 0 |
| Non-intentional firearm discharge (W32–W34) | 4 | 0 | 0 | 0 | 4 | 0 |

**Table S1. Characteristics of the study population with the frequency of each cause of death in our dataset.**[1]

[1] The percentages for the deaths from September 1 2004 to August 31 2013 were obtained by dividing N by the population in September 2004.

| | Type of dementia | | | | All-cause mortality | All-cause mortality except dementia | All-cause mortality except dementia & cerebrovascular diseases |
|---|---|---|---|---|---|---|---|
| | Any type | Alzheimer's disease | Vascular dementia | Unspecified dementia | | | |
| **Total** | | | | | | | |
| Estimate (% points) | -0.38 | -0.06 | -0.06 | -0.14 | -1.23 | -0.73 | -0.52 |
| 95% CI | [-0.68, -0.08] | [-0.19, 0.07] | [-0.25, 0.13] | [-0.39, 0.11] | [-2.35, -0.11] | [-1.61, 0.15] | [-1.49, 0.46] |
| p-value | 0.012 | 0.344 | 0.548 | 0.274 | 0.030 | 0.108 | 0.305 |
| Relative effect (%) | -4.8 | -2.58 | -3.67 | -3.71 | -2.38 | -1.67 | -1.29 |
| Bandwidth (months) | 15.3 | 25.5 | 19.4 | 15.6 | 17.1 | 20.1 | 18.3 |
| Bound on $d^2y/dx^2$ | 0.0037 | 0.0005 | 0.0009 | 0.0026 | 0.0095 | 0.0058 | 0.0067 |
| **Women** | | | | | | | |
| Estimate (% points) | -0.62 | -0.10 | -0.11 | -0.17 | -1.41 | -0.8 | -0.63 |
| 95% CI | [-1.06, -0.19] | [-0.34, 0.14] | [-0.25, 0.03] | [-0.52, 0.18] | [-2.34, -0.48] | [-1.56, -0.04] | [-1.33, 0.07] |
| p-value | 0.004 | 0.415 | 0.144 | 0.345 | 0.002 | 0.036 | 0.083 |
| Relative effect (%) | -6.95 | -3.15 | -6.51 | -4.13 | -2.99 | -2.08 | -1.8 |
| Bandwidth (months) | 13.8 | 23.9 | 22.5 | 13.9 | 18.2 | 20.6 | 21.6 |
| Bound on $d^2y/dx^2$ | 0.0061 | 0.0008 | 0.0009 | 0.0045 | 0.0094 | 0.0062 | 0.0050 |
| **Men** | | | | | | | |
| Estimate (% points) | -0.11 | -0.03 | 0.05 | -0.11 | -0.68 | -0.56 | -0.20 |
| 95% CI | [-0.51, 0.28] | [-0.2, 0.14] | [-0.29, 0.38] | [-0.39, 0.18] | [-2.37, 1.01] | [-1.95, 0.83] | [-1.84, 1.43] |
| p-value | 0.574 | 0.715 | 0.793 | 0.469 | 0.433 | 0.435 | 0.807 |
| Relative effect (%) | -1.63 | -1.61 | 3.29 | -2.96 | -1.18 | -1.11 | -0.44 |
| Bandwidth (months) | 24.1 | 23.9 | 20.3 | 17.4 | 18.7 | 18.4 | 16.4 |
| Bound on $d^2y/dx^2$ | 0.0013 | 0.0008 | 0.0011 | 0.0022 | 0.0085 | 0.0080 | 0.0098 |

**Table S2. Effect estimates of being eligible for herpes zoster vaccination on deaths due to dementia, deaths due to each type of dementia, all-cause mortality except deaths due to dementia, and all-cause mortality except deaths due to dementia or cerebrovascular diseases.**[1]

[1] The bandwidth is the window (in months) in participants' date of birth that is drawn around the September 2 1933 eligibility threshold. We used mean squared error-optimal bandwidths. Abbreviations: CI = Confidence Interval, $d^2y/dx^2$ = second derivative of the conditional mean function of the outcome.

| | Type of dementia | | | | All-cause mortality |
|---|---|---|---|---|---|
| | Any type | Alzheimer's disease | Vascular dementia | Unspecified dementia | |
| Difference by sex (% points) | 0.51 | 0.07 | 0.15 | 0.06 | 0.72 |
| 95% CI | [0.11, 0.91] | [-0.14, 0.28] | [-0.03, 0.34] | [-0.26, 0.39] | [-0.30, 1.75] |
| p-value | 0.014 | 0.521 | 0.105 | 0.690 | 0.165 |
| Bandwidth (months) | W: 13.8, M: 24.1 | W: 23.9, M: 23.9 | W: 22.5, M: 20.3 | W: 13.9, M: 17.4 | W: 18.1, M:17.6 |

**Table S3. Difference between women and men in the effect of being eligible for herpes zoster vaccination on deaths due to dementia and all-cause mortality.** The intent-to-treat (ITT) effect refers to the estimated effect of being eligible for the herpes zoster vaccine. The reference group is women when calculating the difference in the ITT. The difference in the ITT by sex is estimated by estimating the following model:

$$Y_{ts} = \alpha + \beta_1 \cdot (MOB_s - c_0) + \beta_2 D_{ts} + \beta_3 MALE_t + \beta_4 D_{ts} \cdot (MOB_s - c_0) + \beta_5 D_{ts} \cdot MALE_t + \beta_6 \cdot (MOB_s - c_0) \cdot MALE_t + \beta_7 D_{ts} \cdot (MOB_s - c_0) \cdot MALE_t + \epsilon_{ts}$$

where t indexes the month of birth and s indexes the gender. Y is the number of deaths during the follow-up time (i.e., September 1, 2013 – August 31, 2022) divided by the size of the population at the beginning of the follow-up time. The binary variable D indicates eligibility for the herpes zoster vaccine (it is, thus, equal to one after the cutoff date of September 2 1933). The binary variable MALE is equal to one for male month-of-birth observations. The term (WOB-$C_0$) indicates that the month-of-birth observations are centered around the September 2 1933 date-of-birth eligibility date. The interaction term D*(WOB-$C_0$) allows for the slope of the regression line to differ on either side of the date-of-birth eligibility threshold. Adding the terms (WOB-$C_0$)*MALE and D*(WOB-$C_0$)*MALE allows the slopes to vary by gender. The parameter $\beta_5$ identifies the gender difference in the effect of eligibility for the vaccine on the outcome; this is the parameter reported in this table. Consistent with our primary regression discontinuity models shown in the main manuscript, we used the mean squared-error optimal bandwidth sizes from the respective regression model (without interaction terms by gender) in our primary analysis and estimated the effect differences by gender via triangularly weighted least-squared regressions.

Abbreviations: CI=confidence interval; W=women; M=men.

| | Total | Women | Men |
|---|---|---|---|
| Estimate (% points) | -0.28 | -0.67 | -0.02 |
| 95% CI | [-0.49, -0.07] | [-0.98, -0.35] | [-0.27, 0.23] |
| p-value | 0.010 | <0.001 | 0.865 |
| Relative effect (%) | -4.19 | -9.1 | -0.35 |
| Bandwidth (months) | 15.3 | 13.8 | 24.1 |

**Table S4. Effect estimates of being eligible for herpes zoster vaccination on deaths due to dementia as estimated by cause-specific Cox regressions.**[1]

[1] The bandwidth is the window (in months) in participants' date of birth that is drawn around the September 2 1933 eligibility threshold. We used mean squared error-optimal bandwidths from our main regression discontinuity model (these are detailed in Table S2).

Abbreviations: CI = Confidence Interval.
